# Timing matters: Detection of clonal hematopoiesis and its association with adverse outcomes in heart transplant recipients

**DOI:** 10.1101/2025.10.28.25338307

**Authors:** Tafsia Hussain, Darshan H. Brahmbhatt, Fernando Luis Scolari, Sagi Abelson, John E. Dick, Filio Billia

## Abstract

Clonal hematopoiesis (CH) promotes inflammation and is associated with the development of cardiovascular disease. Previous studies assessing CH mutations in orthotopic heart transplant (OHT) recipients have revealed inconsistent findings, likely due to small sample size and differing sample collection time. In this study, we investigated the association between CH and post-transplant outcomes with a more consistent sample collection window. This retrospective study included 209 patients who underwent OHT between 2015 and 2022. Targeted sequencing detected CH mutations from samples obtained within a window of six months before or after transplantation. Clinical data were collected from the electronic medical record. Patients undergoing OHT had a median age of 53 years, and 27% were female. CH-associated mutations with a variant allele frequency (VAF) greater than 2% were detected in 29 patients (13.9%). The commonly mutated genes included DNMT3A, TET2, and ASXL1. CH mutations were associated with an increased risk of antibody-mediated rejection (AMR) (HR 2.42, 95% CI 1.07-5.47, p=0.033), but without detected differences in mortality or cardiac allograft vasculopathy (CAV). CH mutations detected at the time of transplant were associated with clinically significant AMR. Sample analysis at the time of transplant provides the clearest association between CH mutations and outcomes in OHT.

## INTRODUCTION

Orthotopic heart transplant (OHT) is the definitive treatment for patients with end-stage heart failure (HF) and refractory symptoms that cannot be managed with optimal medical therapy. The annual number of OHTs performed continues to grow with improvement in outcomes due to enhancements in donor and recipient management.^1^ In recent years, the 1-year post-transplant survival has reached 92%^1^, reflecting improvements in perioperative care and immunosuppressive strategies. However, early survival is still influenced complications such as the development of primary graft dysfunction, post-operative infection and acute cellular rejection, whereas late phase challenges including the development of malignancy, infection, cardiac allograft vasculopathy (CAV) and other complications related to long-term use of immunosuppression.^2–5^

A number of studies have investigated the role of clonal hematopoiesis (CH) in altering the prognosis in patients with HF^6–8^, and more specifically after OHT.^9–11^ CH is an emerging phenomenon linked to ageing, inflammation and cardiovascular disease that has been shown to increase the risk of developing coronary artery disease. CH may also have a role in altering the course of the post-transplant immune response and the development of graft dysfunction after OHT.^9^

CH is a clonal expansion of hematopoietic stem and progenitor cells that have acquired specific somatic mutations.^12–15^ Studies over the past decade have found that the proportion of mutant hematopoietic stem cells increase with age.^15^ Some CH mutations drive increased myeloid differentiation and are linked to inflammation and cardiovascular diseases. CH mutations have also been observed to create an inflammatory environment within the body, through the increased release of different cytokines and chemokines.^12,16–18^ Interestingly, the production of several cytokines and chemokines have been shown to be related to graft tolerance, rejection episodes and ischemic-reperfusion injury.^19–21^.

Studies of CH in patients after OHT have provided contradictory results with regards to association of CH and post-transplant adverse outcomes. The first report in this field, observed CH mutations in 20% of the cohort with the key finding that only OHT recipients with CH developed severe CAV.^9^ In addition, patients with CH had a higher mortality compared to those without CH. However, other studies reported inconsistent results regarding the association between CH and post-transplant adverse outcomes.^10,11^ These opposing findings are likely due to variations in study design and the unknown effects of immunosuppression on post-OHT clonal expansion. Therefore, this current study specifically investigates the association between CH at the time of transplant and adverse outcomes in OHT recipients.

## MATERIALS AND METHODS

### Study population

This retrospective study followed the Declaration of Helsinki and was approved by the University Health Network (UHN) Research Ethics Board (#22-5005). A total of 264 adult OHT recipients from 2015 to 2022 were screened for this study. Patients who were adults (≥18 years) at the time of transplant, provided informed consent to the Peter Munk Cardiac Centre (PMCC) Biobank and had a biospecimen collected within 6 months prior to or after the transplant procedure were included in this study. These patients’ blood sample underwent sequencing to detect CH as previously described.^9^

### Data collection

Clinical data for each OHT recipient was collected from their electronic health record. All the clinical data was organized and stored in a Research Electronic Data Capture (REDCap) database. Baseline characteristics were collected which included demographic data, comorbidities, and transplant crossmatch data. Laboratory assessments as close as possible to the collection date of the biospecimen were recorded. Medications including immunosuppressive therapy at the time discharge and one-year follow up were documented. In addition, early post-transplant outcomes such as primary graft dysfunction, use of temporary mechanical circulatory support (MCS), and renal replacement therapy were documented. The first occurrence of each major outcome including CAV, acute cellular rejection (ACR), antibody-mediated rejection (AMR), *de novo* malignancy, various infections and mortality was recorded. For outcomes graded by severity, the highest documented grade was documented. Cumulative occurrences of ACR in the first year were also counted.

### Follow-up and outcomes

Each patient was followed from the time of their transplant procedure to their last visit to the Ajmera Transplant Centre or the time of death. ACR and AMR were confirmed through endomyocardial biopsies and graded based on the standards set by the International Society for Heart and Lung Transplantation (ISHLT).^22^ The first incidence of ACR grade 2R or 3R were noted. Similarly, patients who developed AMR grade 1-3 were recorded. This grade was determined by histological features and immunohistochemical staining for C4d. Serological evidence of circulating donor-specific antibodies (DSA) were also assessed to assist in determining AMR. CAV was diagnosed and confirmed through routine coronary angiography and graded based on the standards outlined by ISHLT.^23^ *De novo* malignancy was diagnosed through tissue biopsy conducted as per hospital protocols.

Blood cultures, urinalysis, lactate levels, C-reactive protein (CRP), and clinical notes were employed to define a positive sepsis event. PCR testing of blood samples identified OHT recipients with cytomegalovirus (CMV) and Epstein-Barr virus (EBV) viremia. Fungal infections were confirmed via positive fungal cultures, while Candida species were identified through positive blood culture results. Non-tuberculous mycobacteria (NTM) infections were diagnosed using Acid-Fast Bacilli (AFB) smears, and Clostridioides difficile (C.difficile) infections were confirmed with a toxin assay. Chest radiographs combined with blood cultures were utilized to confirm pneumonia, while urinalysis was used to diagnose urinary tract infections (UTIs). Additionally, infections such as COVID-19 or Escherichia coli (E. Coli) were documented, if recorded in the patient’s electronic health records. Any cause of mortality was documented.

### Genetic sequencing procedure

Biospecimens were collected from OHT recipients with consent at the Ajmera Transplant Centre and stored in the Peter Munk Cardiac Centre (PMCC) biobank. Biospecimens collected within ±6 months of the transplant date were sequenced. A patient with CH mutations was defined as having a VAF equal to or more than 2%. Variant calling steps were carried out as described in Scolari et al.^9^ In summary, the single molecule Molecular Inversion Probes (smMIP) technique was employed. A genomic library was constructed with the genes of interest. The Illumina Novaseq Platform then generated pair-end 150 base pair sequences. These sequences were produced according to the genes which have been found to be mutated in myeloid neoplasms. Additional details are provided in online supplementary *Appendix S1*.

### Statistical analysis

Continuous variables are presented as median and interquartile range (IQR). Categorical variables are displayed as counts and percentages. Using the Shapiro-Wilk test, all variables were tested for normality. Comparisons between the no CH and CH groups were conducted using the Wilcoxon rank sum test for continuous variables. For categorical variables, the Chi-square test or Fisher’s exact test was employed. Comparable analyses were performed on outcome-related variables to compare individuals with DNMT3A, TET2, or ASXL1 mutations to those without CH mutations. Similar additional analyses were conducted to compare the patients who were sequenced to those who were not sequenced to detect CH.

A multivariable logistic regression model was performed with a 95% confidence interval to evaluate the association between CH mutations or CH-related DNMT3A, TET2, and ASXL1 mutations and major adverse outcomes. A univariable and multivariable Cox proportional hazards model was computed for CAV, AMR, ACR, malignancy, and mortality outcomes. The Cox proportional hazards models were adjusted based on covariates including age, sex, body mass index (BMI), ischemic etiology, diabetes, hypertension, dyslipidemia previous myocardial infarction and pre-transplant intracardiac device implantation. Survival function according to CH status was represented using the Kaplan-Meier curve. Fine-Gray subdistribution hazard model was used to estimate the cumulative incidence of CAV grade 1-3 and CAV 2-3, with death being a competing event. All statistical tests were 2-tailed and a statistical significance of .05 was used. All analyses were performed using R 4.3.2.

## RESULTS

### Baseline characteristics

A total of 264 adult patients who underwent an OHT at the Ajmera Transplant Centre (University Health Network) between 2015 and 2022 were initially identified (Figure 1). Twelve patients were excluded due to lack of consent to the PMCC biobank. Of the remaining 252 patients, 43 were excluded as no biospecimen was collected within six months before or after transplant. The final study cohort includes 209 patients whose samples were then sequenced.

**Figure 1.**
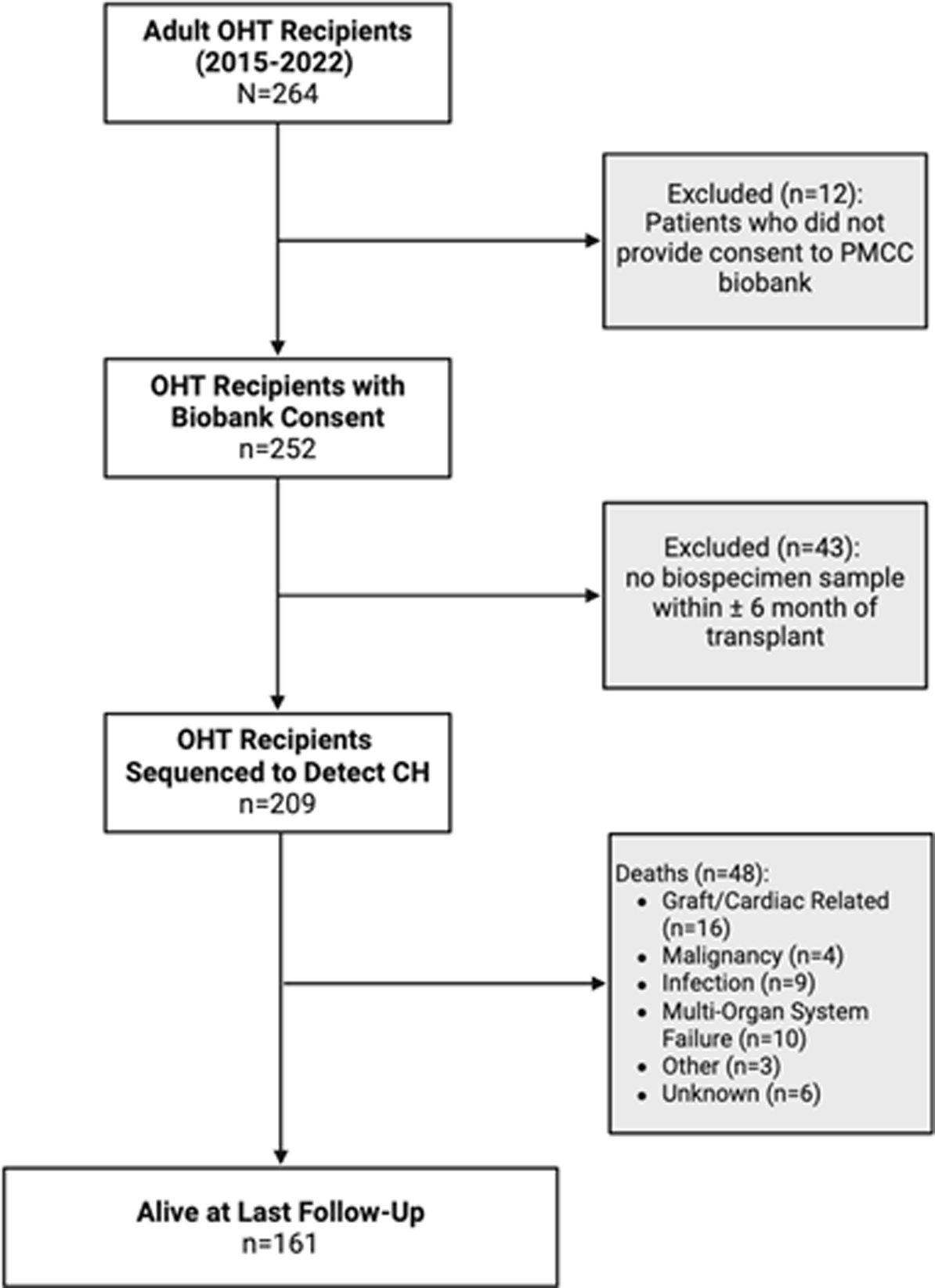
Flowchart of the study population. A total of 264 adult patients who received a heart transplant between 2015 and 2022 were initially identified. Twelve patients who did not consent to the biobank were excluded. Of the remaining 252 patients, 43 were excluded due to lack of a biospecimen collected within six months before or after transplant. The final cohort included 209 patients whose samples were sequenced. Details of the 43 excluded patients who did not have biospecimen available are provided in the Supplementary Table 1 and 2. OHT, orthotopic heart transplant.

The study cohort of 209 OHT patients had a median follow-up time post-OHT of 5.1 years (IQR: 3.3 years). The median age was 53 years (IQR 41-60) (Table 1). The most common comorbidities in the study cohort were chronic kidney disease (CKD) (n=59, 28%), dyslipidemia (n=54, 26%), hypertension (n=51, 24%), atrial fibrillation (n=59, 28%) and diabetes mellitus (n=42, 20%). In this cohort, 32% of patients received an implantation of left ventricular assist device (LVAD) as bridged to transplant. The most common etiology of HF was non-ischemic (67%), followed by ischemic (21%) and congenital heart disease (13%). Table 1 provides additional details of the patient’s baseline characteristics.

### Prevalence and distribution of CH mutations

The median interval between sample collection and transplant was 1 day. There were 29 patients out of 209 who had CH mutations with a VAF over 2%. This was a prevalence of 13.9% (Figure 2a). There were 18 CH genes found to be mutated, and the top three commonly mutated genes included DNMT3A (8 patients), TET2 (5 patients), and ASXL1 (4 patients) (Figure 2b). The largest proportion of patients with CH mutations (12%) were between 51–60 years of age (Figure 2c). Comparable proportions were noted in female (44%) and male (40%) OHT recipients with CH mutations (Figure 2d).

**Figure 2.**
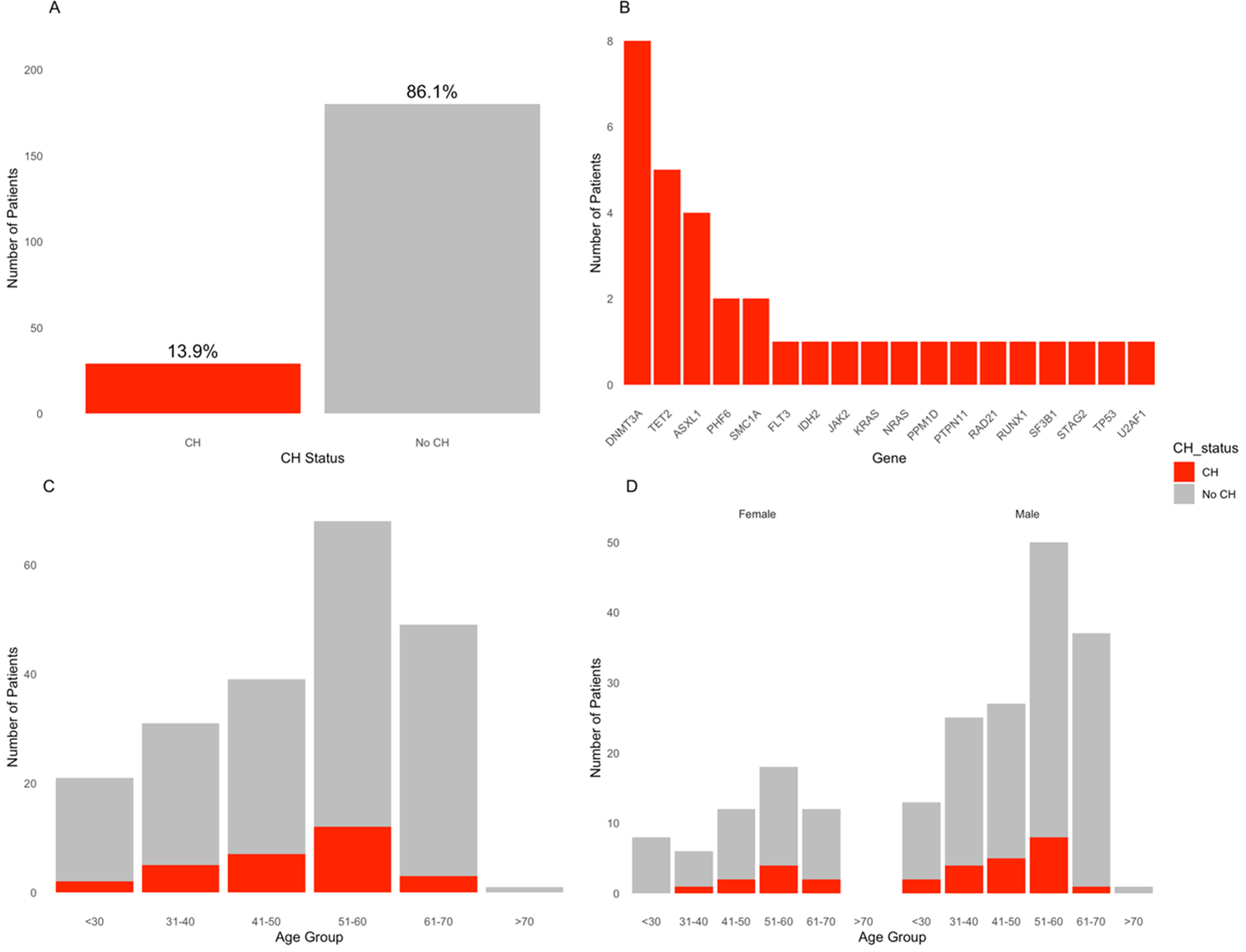
Prevalence and distribution of CH mutations with VAF> 2% in OHT recipients. Panel A shows the prevalence of CH. Panel B presents the distribution of specific mutations among OHT recipients with CH. Panel C displays the distribution of CH by age groups. Panel D further breaks down this distribution by age groups among female and male OHT recipients. VAF, variant allele frequency; CH, clonal hematopoiesis, OHT, orthotopic heart transplant.

The median duration of follow-up was 5.1 years (IQR 3.2 years) for patients without CH, compared with 5.2 years (IQR 3.6 years) for those with CH. Age was not statistically significant between CH and no CH groups (51 vs. 53, p=0.5) (Table 1). When comparing baseline characteristics between patients with and without CH mutations, no significant differences were observed in BMI, hypertension, dyslipidemia, diabetes mellitus, smoking history, CKD, prior myocardial infarction, cerebral vascular accident (CVA)/transient ischemic attack (TIA) and history of alcohol abuse. Similarly, this was also observed in surgical interventions before transplant such as percutaneous coronary intervention (PCI), coronary artery bypass graft surgery (CABG) and implantation of LVAD. In the CH group, most patients had an ICD in place before their transplant (n=12, 52%) and had a non-ischemic heart failure etiology (n=19, 66%) although these were not significantly different.

### Association between CH mutations and major adverse outcomes

OHT recipients with CH mutations had statistically significantly increased AMR Grade 1-3 (31% vs 16%, p=0.043) (Table 2), with a multivariate hazards ratio (HR) of 2.42 (CI 95% 1.07-5.4, p=0.033) (Table 3). Given the established association of AMR with both CAV and mortality, we subsequently evaluated these outcomes within our cohort. There was no association between all-cause mortality and CH. Six (21%) patients with CH patients died post-transplant, while 42 (23%) recipients with no CH died (p=0.8) (Table 2). Early mortality was seen in OHT recipients with CH and reached to about 20.6% cumulative incidence at 2 years followed by a plateau (Supplementary Figure 2a). In a time-to-event analysis, there was no difference in survival between those who had CH and those who didn’t (p=0.8) (Figure 3). In addition, there was no association between moderate to severe CAV (defined as CAV grade ≥ 2) to those who had CH mutations (6.9% vs. 5.0%, p=0.7) (Table 2). This was also observed in patients had any CAV (Grade 1-3) (66% vs. 57%, p=0.4). With a competing risk analysis as death being a competing event, there was no significant difference between CAV and CH (p=0.36) (Figure 4a). Similarly, this was also seen in moderate to severe CAV (p=0.65) (Figure 4b).

**Figure 3.**
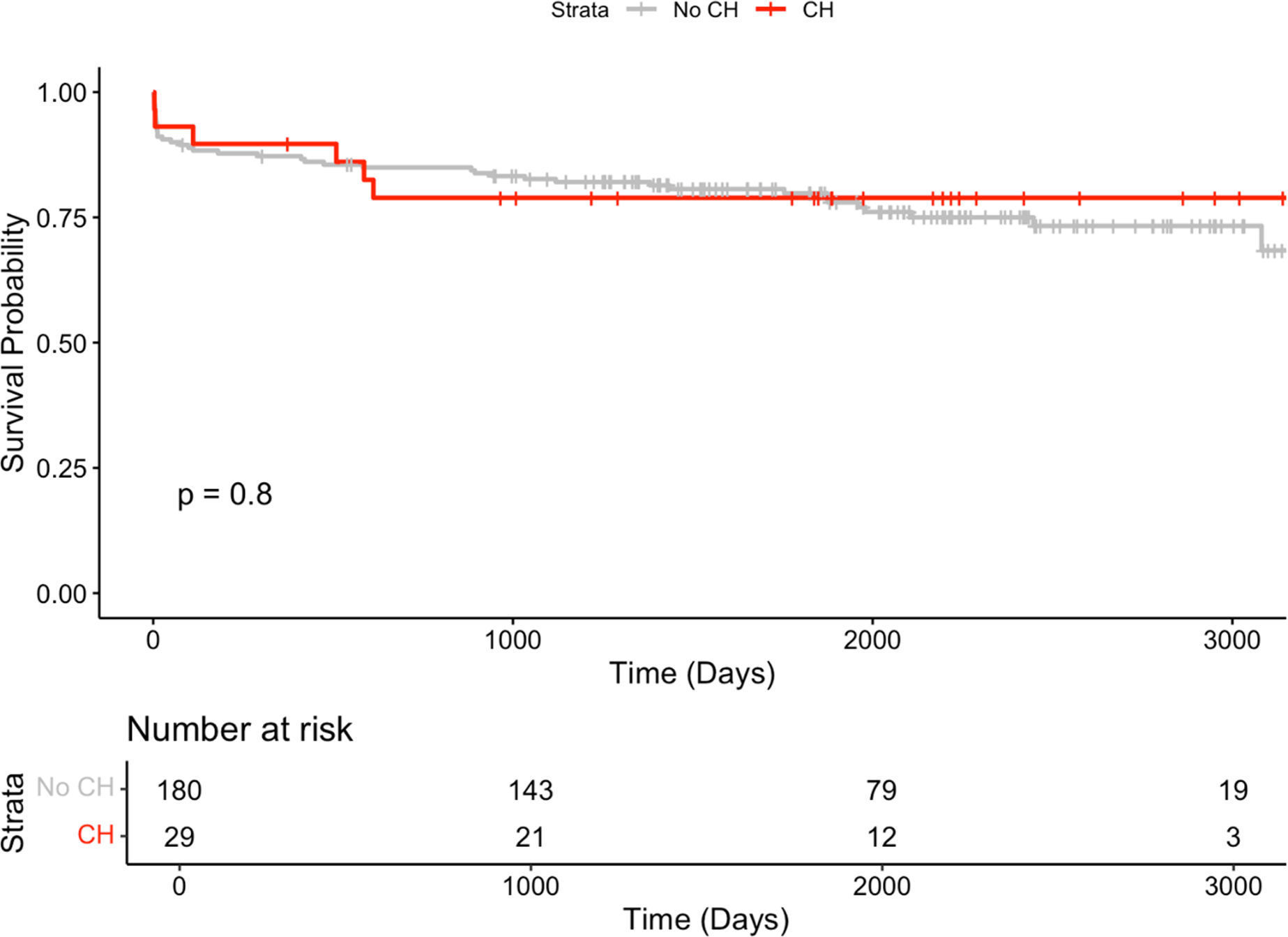
Kaplan-Meier survival curve of OHT recipients stratified by CH status with a VAF≥ 2%. Time represents days post-heart transplant. CH, clonal hematopoiesis; OHT, orthotopic heart transplant.

**Figure 4.**
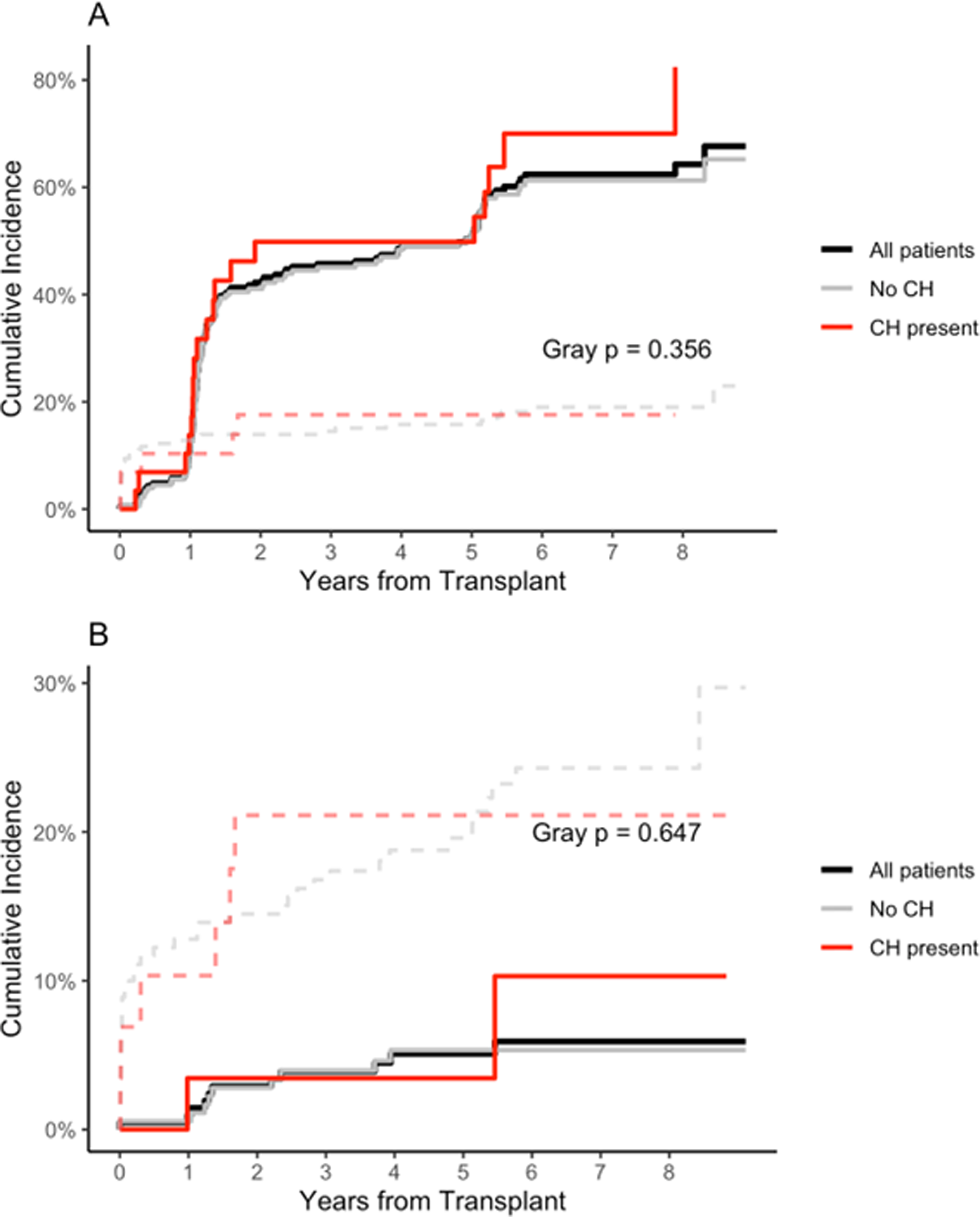
Competing-risk cumulative incidence of CAV after OHT, stratified by CH status with a VAF ≥ 2%. Panel A represents competing-risk cumulative-incidence function for grade 1–3 CAV, death treated as a competing event. Panel B presents the competing-risk cumulative-incidence function for grade ≥ 2 CAV with death as the competing event. Solid curves represent incidence of CAV; dashed curves track the competing event death without prior CAV. Gray-test P values compare CHDpositive versus CHDnegative sub-distributions. VAF, variant allele frequency; CH, clonal hematopoiesis, OHT, orthotopic heart transplant, CAV, cardiac allograft vasculopathy.

In addition, no associations were found between CH mutations and ACR Grade 2R/3R, de novo donor-specific antibodies (DSA), malignancy, sepsis, CMV viremia, EBV viremia, or other infections (Table 2, Supplemental Table 7). Early outcomes post-surgery including primary graft dysfunction, temporary MCS and temporary dialysis were not statistically significant (Table 2).

## DISCUSSION

The results of this study demonstrate a significant association between CH mutations assessed at the time of OHT and AMR. This is the first study that assesses CH driver mutations within 6 months of the time of transplant, thus avoiding any concerns of survivor selection bias encountered when patients are recruited post-transplant, or if the assessment of CH is undertaken many years before transplant, both of which may alter true prevalence. Moreover, if CH is assessed many months after OHT, exposure to immunosuppression may also influence its detection and prevalence.

### Inflammation implicated as the link from CH to development of AMR

AMR is one of the leading causes of graft failure and death in OHT recipients, as it has shown to increase the risk of CAV development.^24–26^ The role of inflammation is well documented in the process of AMR.^27–29^ There are a number of immunological and inflammatory factors which are activated by a number of triggers, including donor organ arrest and procurement, graft ischemic time and reperfusion post-transplant.^30,31^ Given that CH mutations has been shown to augment the body’s inflammatory response and linked to adverse cardiovascular outcomes, it is not a great leap to ascribe elevated post-transplant inflammation effects to related to the presence of CH.^12,17,18,32,33^

Several mouse model studies have investigated common CH mutations which have shown to be linked to increased production of pro-inflammatory mediators. Mice with DNMT3A deficient mast cells were observed to have a rise in IL-6, TNF-a and IL-13 production.^34^ Likewise, under inflammatory stress, TET2-knockout mice showed higher levels of several inflammatory cytokines and chemokines. Notably, there was an enhanced production in IL-6, Ccl2, Ccl4, TNF--a and CXCL9 after the lipopolysaccharide treatment.^35^ These findings suggest CH mutations create a pro-immune milieu in which HLA expression is upregulated, ultimately promoting the DSA response leading to rejection.^36^ This may provide a plausible mechanistic link between CH and AMR. OHT recipients who have CH mutations at the time of transplantation may enter the postoperative period with a heightened innate immune system, driving AMR.

In this study, the association of CH mutations and CAV is not seen, however, given the known link between AMR and CAV, with a longer follow up time an association may have emerged. It is known in the literature, inflammation in the longer term also leads to progression of CAV.^37–39^ It has also been shown CH is associated in the progression of ischemic heart disease^33^, which has been shown to have similar inflammatory signalling pathways to allograft vasculopathy.^40^ This may suggest CAV can be plausibly accelerated in the presence of CH mutations. It remains to be seen what effect treatments for AMR prevention has on CH in patient’s post-OHT to better understand the link between CH and inflammation after transplant.

### Timing of sample and effect on outcomes

This is the fourth study that has investigated the association of CH with outcomes post-OHT. The first study, from our own group, Scolari et al, was a convenience sample of 127 OHT patients with samples acquired pre- and post-OHT (68% with samples acquired a median 200 days pre-transplant and 32% patients were enrolled post-transplant, with samples collected a median of 114 days after OHT).^9^ This showed an association between recipient CH and both the development of CAV and mortality. Notably, 21 patients from the Scolari et al. cohort were not included in the present study because their transplant occurred prior to 2015, or their sample was collected outside the ±6-month post-OHT window. Among those excluded, 3 developed CAV and 6 died, which may account for the differences in outcomes observed between the two studies.

Further studies by Simitsis et al^11^ (which enrolled patients 8.5 years after transplant) and Amancherla et al^10^ (a pooled analysis of patients from Vanderbilt University Medical Center, VUMC, sequenced 2.3 years before transplant and Columbia University Irving Medical Center (CUIMC) patients who had biospecimens taken at time of transplant) did not show any association between CH and post-OHT outcomes. The current study shows significant associations of CH with AMR.

The estimated CH prevalence is closely related to the timing of sample acquisition when considering time of transplant. Between the two cohorts, the Amancherla et al study had a 14.9% CH prevalence^10^, similar to this current study of 13.9% where both acquired samples close to the time of OHT. Finally, the Simitsis et al study showed a prevalence of 31.6%, but these were acquired a median of 8.5 years after OHT.^11^ The justification for using such a wide range of sample acquisition timings is based on two studies which showed that mutations did not appear to change very much over the course of three and six years.^41,42^ However, these studies were conducted in healthy elderly patients in their eighth decade, not affected by cardiovascular disease. Moreover, this overlooks the high-risk rapidly expanding VAF clone which is more likely to be pathogenic.^41^ For this reason, we argue that the results of the VUMC cohort are likely to underestimate CH at the time of transplant, in fact the CH prevalence is significantly different between the VUMC and CUIMC cohorts (82/609 vs 35/178; Chi squared p=0.041) (Supplementary Figure 1). There is a selection bias where only 301 of 609 were included from the VUMC site. The findings of the Simitsis et al study also warrant concern for selection bias. As patients were enrolled 8.5 years after transplant, it is likely that a number of patients will have had CAV or AMR and died before sampling would have occurred. When timing of sample acquisition is not standardized, results can be confusing to discern, as seen in a recent biobank study of CH in all solid organ transplant where samples were collected at the time of biobank enrollment, not explicitly reporting the time between transplantation and enrolment.^43^

### Limitations

In this retrospective study, various limitations are present. This is a single centre study which may introduce potential bias. A multicenter approach would further improve data applicability, drawing on diverse patient demographics. The second limitation is the short follow-up period after transplant with an average follow-up time of about 5 years. A longer follow-up time may be needed to demonstrate the association of CH and additional adverse outcomes post-transplant. In addition, a subset of eligible patients in the biobank between 2015-2022 lacked a sample collected within the ±6-month window from the transplant date. Analyses suggests that exclusion of these patients is unlikely to have materially influenced the overall conclusions. Finally, the study is under-powered for outcomes. Only with a larger cohort it will be able to confirm or refute the observed effect sizes.

### Future directions

Future work should extend beyond our single-time-point analysis by prospectively following OHT recipients with serial sample collection, and collecting data on key events such as CAV, AMR, and graft failure. In addition, with multiple timepoints, clone size and specific driver mutations may provide insights into which mutations confer the greatest risk and may reveal therapeutic treatment windows. Multi-centre cohorts are needed to test whether immunosuppressive regimens, lipid-lowering agents, or anti-inflammatory therapies modify CH. These future directions could significantly advance our understanding and management of CH in transplant medicine.

## CONCLUSIONS

This study examined the prevalence of CH with a VAF over 2% in OHT recipients and its association with post-transplant adverse outcomes. There was an 13.9% prevalence of CH with biospecimens collected at the time of transplant. The presence of CH at transplant was significantly associated with an increased risk of AMR. These findings emphasize the importance of when CH is diagnosed, as it may provide insight into a recipient’s risk for adverse post-transplant outcomes in the longer term.

## Data Availability

All datat produced in the presented are contained in the manuscript.

## Abbreviations

ACR: Acute cellular rejection
AMR: Antibody-mediated rejection
ASXL1: Additional sex combs-Like 1
BMI: Body mass index
CABG: Coronary artery bypass grafting
CAV: Cardiac allograft vasculopathy
CH: Clonal hematopoiesis
CKD: Chronic kidney disease
C.difficile: Clostridioides difficile
CMV: Cytomegalovirus
CRT-D: Cardiac resynchronization therapy with defibrillator
CRT-P: Cardiac resynchronization therapy with pacemaker
DNMT3A: DNA methyltransferase 3 alpha
DSA: Donor-specific antibodies
EMB: Endomyocardial biopsy
HF: Heart failure
HR: Hazard ratio
ICD: Implantable cardioverter defibrillator
OHT: Orthotopic heart transplant
PCI: Percutaneous coronary intervention
PPM: Permanent pacemaker
MCS: Mechanical circulatory support
TET2: Ten-eleven translocation 2
VAD: Ventricular assist devices
VAF: Variant Allele Frequency

## ACKNOWLEDGEMENTS

Conceptualization: Tafsia Hussain, Darshan H. Brahmbhatt, Fernando Luis Scolari, John E. Dick, Sagi Abelson, Filio Billia

Study design: Tafsia Hussain, Darshan H. Brahmbhatt, Fernando Luis Scolari, John E. Dick, Sagi Abelson, Filio Billia

Data collection: Tafsia Hussain

Data analysis: Tafsia Hussain, Darshan H. Brahmbhatt, Fernando Luis Scolari

Data interpretation: Tafsia Hussain, Darshan H. Brahmbhatt, Fernando Luis Scolari, Filio Billia

Writing original draft: Tafsia Hussain, Darshan H. Brahmbhatt

Review and editing: Tafsia Hussain, Darshan H. Brahmbhatt, Fernando Luis Scolari, John E. Dick, Sagi Abelson, Filio Billia

Funding acquisition: Filio Billia and John E. Dick

Special thank you to Drs. Steven Chan and Robert Vanner for their expertise and feedback.

## Data availability

Once the study is published the datasets, including the redacted study protocol, redacted statistical analysis plan, and participant data supporting the results reported in this article, will be available to researchers who provide a methodologically sound proposal, at the discretion of the institutional Research Ethics Board. The data will be provided after its de-identification, in compliance with applicable privacy laws, data protection, and requirements for consent and anonymisation.

