## supplemental material for "Timing matters: Detection of clonal hematopoiesis and its association with adverse outcomes in heart transplant recipients"

**SUPPLEMENTARY APPENDIX**

##### Supplementary Materials and Methods

**Data Collection, Follow-Up and Outcomes**

We collected pre-transplant data, intraoperative to post-transplant outcomes. For all patients, age, sex, height, weight, BMI were collected. In addition, comorbidities recorded include dyslipidemia, diabetes, smoking and alcohol abuse. Specific to heart transplant recipients, heart failure etiology, history of hypertension, atrial fibrillation, cerebral vascular accident (CVA), transient ischemic attack (TIA), peripheral vascular disease, myocardial infarction, implantation of intracardiac device or cardiac surgery were noted.

Donor information such as blood type, sex, CMV status, and EBV status were also recorded. Other intraoperative variables which were noted included CMV status and EBV status of the recipient, induction therapy, results of the crossmatch test, pre-transplant donor specific antibodies (DSA), Class I/Class II HLA test, ischemic time, bypass time.

**Biospecimen Collection**

With the patient's consent, all biospecimen were collected at UHN and stored in a biobank. Peripheral blood samples were collected from the patients nearest to the procedure date. Biospecimens of heart transplant recipients were stored in the Peter Munk Cardiac Centre (PMCC) Cardiovascular Biobank.

**Genetic Sequencing Procedure**

The genomic libraries were created using the single-molecule Molecular Inversion Probes (smMIP). These libraries included the following genes: ASXL1, BCOR, BRAF, CALR, CBL, CEBPA, DNMT3A, EZH2, FLT3A, GATA1, GATA2, GNAS, IDH1, IDH2, JAK2, KIT, KRAS, MPL, NRAS, PHF6, PPM1D, PTPN11, RAD21, RUNX1, SETBP1, SF3B1, SMC1A, SMC3, SRSF2, STAG2, TET2, TP53, U2AF1, WT1, ZRSR2.

To reduce artifacts and false positives in identifying mutations, samples were sequenced twice, and a series of data processing steps were completed using a computational pipeline developed at our institution (Medeiros JJF, et al. Bioinformatics 2022).

Somatic variant calling for CH was performed using SmMIP-tools, followed by a multi-step filtering process to identify high-confidence mutations. Variants were excluded if they carried sequencing or analytical flags. Only variants with a Combined Annotation Dependent Depletion (CADD) score ≥ 10 were retained. We required acceptable ratios between the number of samples with statistically significant P-values (Bonferroni-corrected ≤ 0.05) and the number of replicates in which the variant passed this threshold. To minimize the inclusion of germline variants, clonal mutations with a minor allele frequency (MAF) ≥ 0.001 were excluded, except for hotspot mutations previously reported in the Catalogue of Somatic Mutations in Cancer (COSMIC). Technical support was further evaluated using the SSCS-derived (SSCS-drop) collapse ratio, and only variants with a value > 1.5 were accepted. Variants failing the P-value criterion were reconsidered if they had both a CADD score ≥ 10 and an SSCS-drop > 2.0. Variants with an SSCS-drop < 1.0–1.2, or between 1.2–1.5 with poor statistical support and COSMIC absence, were also excluded. Furthermore, variants were removed when observed at higher variant allele frequencies (VAF) in other samples within the cohort, particularly if accompanied by low SSCS-drop or poor P-value support. Finally, only variants with a variant allele frequency (VAF) > 2% were included in the analysis.

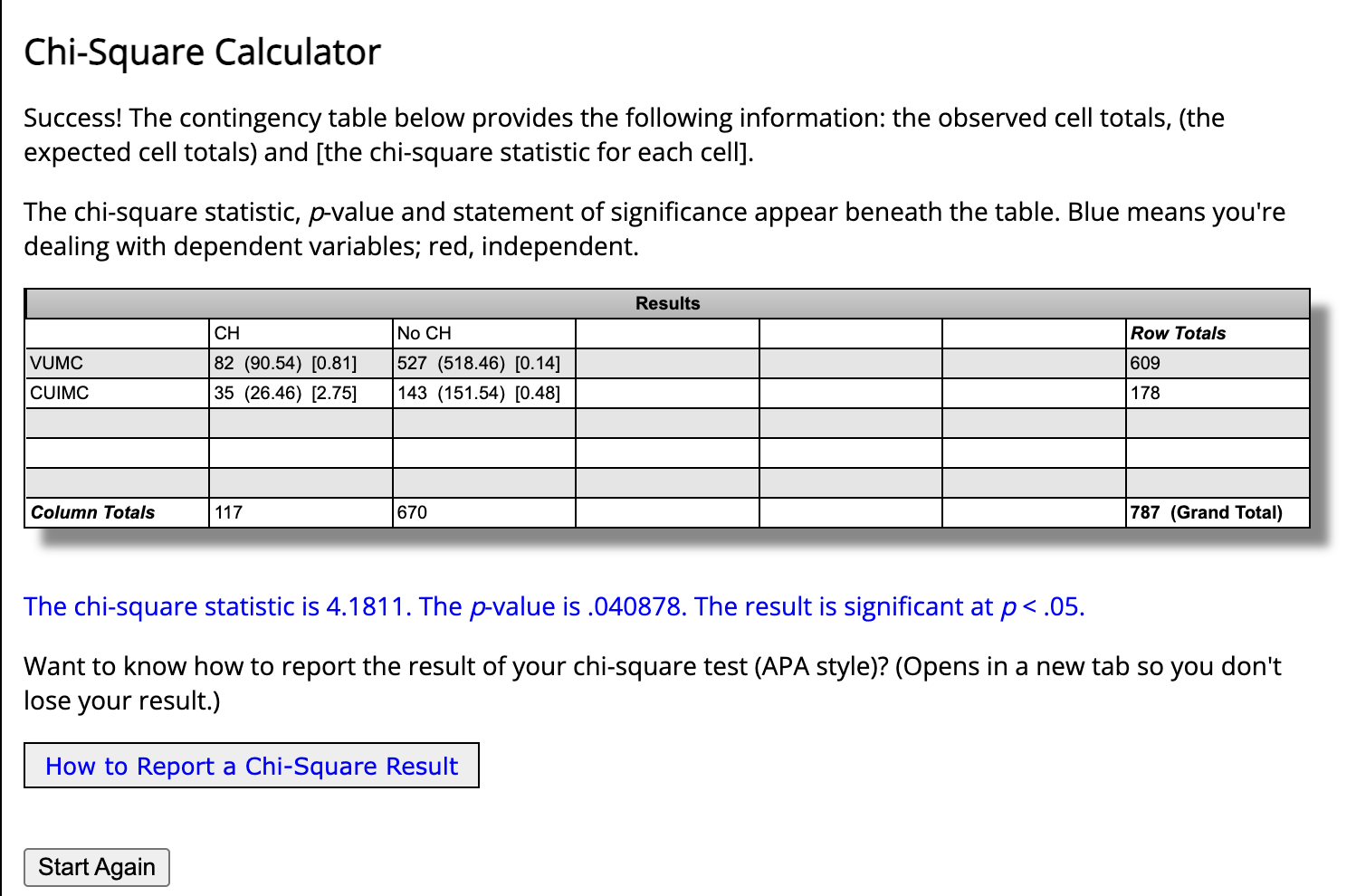

##### **Supplementary Figure 1.**

### Orthotopic Heart Transplant (OHT) Recipients at the Ajmera Transplant Centre (2015-2022)

##### **Supplementary Table 1.** Comparison of baseline characteristics between sequenced and non-sequenced OHT biobank patients.

| **Characteristic** | **All Biobank Patients,**  **N = 252^1^** | **Study Cohort (Sequenced),** n = 209*^1^* | | **Non-sequenced Patients,** n = 43*^1^* | | **p-value***^2^* | |
| --- | --- | --- | --- | --- | --- | --- | --- |
| Age (years) | 53 (40, 60) | | 53 (41, 60) | | 54 (38, 58) | | 0.6 |
| Sex |  |  | |  | | >0.9 | |
| Male-no. (%) | 184 (73%) | 153 (73%) | | 31 (72%) | |  | |
| Female-no. (%) | 68 (27%) | 56 (27%) | | 12 (28%) | |  | |
| BMI (kg/m^2^) | 25.1 (22.4, 28.7) | 25.2 (22.4, 28.9) | | 25.1 (22.2, 28.1) | | 0.6 | |
| History of dyslipidemia-no. (%) | 63 (25%) | 54 (26%) | | 9 (22%) | | 0.6 | |
| History of diabetes mellitus-no.% | 49 (20%) | 42 (20%) | | 7 (17%) | | 0.7 | |
| History of hypertension-no.% | 59 (24%) | 51 (24%) | | 8 (20%) | | 0.5 | |
| History of myocardial infarction-no.% | 41 (16%) | 33 (16%) | | 8 (20%) | | 0.6 | |
| Previous PCI -no.% | 28 (11%) | 24 (11%) | | 4 (9.8%) | | >0.9 | |
| Previous CABG-no.% | 13 (5.2%) | 12 (5.7%) | | 1 (2.4%) | | 0.7 | |
| History of CVA/TIA-no.% | 45 (18%) | 37 (18%) | | 8 (20%) | | 0.8 | |
| History of atrial fibrillation/flutter-no.% | 63 (25%) | 59 (28%) | | 4 (9.8%) | | **0.013** | |
| History of CKD-no.% | 70 (28%) | 59 (28%) | | 11 (27%) | | 0.9 | |
| History of alcohol abuse-no.% | 13 (5.2%) | 10 (4.8%) | | 3 (7.3%) | | 0.4 | |
| History of smoking-no.% | 60 (24%) | 51 (24%) | | 9 (22%) | | 0.2 | |
| Intracardiac device | | | | | | | 0.11 |
| ICD-no.% | 104 (54%) | 81 (50%) | | 23 (70%) | |  | |
| CRT-P-no.% | 4 (2.1%) | 4 (2.5%) | | 0 (0%) | |  | |
| CRT-D-no.% | 86 (44%) | 76 (47%) | | 10 (30%) | |  | |
| Etiology of Heart Failure | | | | | | | 0.6 |
| Ischemic-no.% | 49 (20%) | 43 (21%) | | 6 (14%) | |  | |
| Non-ischemic-no.% | 170 (68%) | 139 (67%) | | 31 (74%) | |  | |
| Congenital-no.% | 32 (13%) | 27 (13%) | | 5 (12%) | |  | |
| ABO Group |  |  | |  | | 0.12 | |
| A-no.% | 98 (39%) | 79 (38%) | | 19 (46%) | |  | |
| B-no.% | 39 (16%) | 29 (14%) | | 10 (24%) | |  | |
| AB-no.% | 13 (5.2%) | 12 (5.7%) | | 1 (2.4%) | |  | |
| O-no.% | 100 (40%) | 89 (43%) | | 11 (27%) | |  | |
| *^1^* Median (Q1, Q3); n (%)  *^2^* Wilcoxon rank sum test; Pearson’s Chi-squared test; Fisher’s exact test  Bolded p-values indicates statistical significance.  BMI, Body mass index; PCI, Percutaneous coronary intervention; CABG, Coronary artery bypass surgery; CVA, cerebral vascular accident; TIA, transient ischemic attack; CKD, chronic kidney disease; ICD, implantable cardioverter-defibrillator; CRT-D, cardiac resynchronization therapy defibrillator; CRT-P, cardiac resynchronization therapy pacemaker; OHT, orthotopic heart transplant | | | | | | | |

##### **Supplementary Table 2.** Comparison of post-transplant adverse outcomes between sequenced and non-sequenced OHT biobank patients.

| **Characteristic** | **All Biobank Patients,**  **N = 252^1^** | **Study Cohort (Sequenced),** n = 209*^1^* | **Non-Sequenced Patients,** n = 43*^1^* | **p-value***^2^* |
| --- | --- | --- | --- | --- |
| Antibody-Mediated Rejection Grade 1-3-no.% | 49 (20%) | 37 (18%) | 12 (28%) | 0.14 |
| CAV Grade |  |  |  | 0.2 |
| CAV 0/1-no.% | 241 (96%) | 198 (95%) | 43 (100%) |  |
| CAV 2/3-no.% | 11 (4.4%) | 11 (5.3%) | 0 (0%) |  |
| Acute cellular rejection (2/3R)-no.% | 147 (59%) | 126 (61%) | 21 (49%) | 0.13 |
| Acute cellular rejection-first-year post-transplant-no.% | 130 (52%) | 113 (55%) | 17 (40%) | 0.063 |
| De novo DSA-no.% | 44 (18%) | 38 (19%) | 6 (14%) | 0.5 |
| Malignancy-no.% | 48 (19%) | 39 (19%) | 9 (21%) | 0.7 |
| Infections |  |  |  |  |
| Any infection-no.% | 201 (80%) | 161 (77%) | 40 (93%) | **0.017** |
| Sepsis-no.% | 53 (21%) | 42 (20%) | 11 (26%) | 0.4 |
| CMV Viremia-no.% | 54 (21%) | 44 (21%) | 10 (23%) | 0.7 |
| EBV Viremia-no.% | 28 (11%) | 20 (9.6%) | 8 (19%) | 0.11 |
| Candida-no.% | 27 (11%) | 19 (9.1%) | 8 (19%) | 0.10 |
| Fungal-no.% | 25 (9.9%) | 18 (8.6%) | 7 (16%) | 0.2 |
| UTI-no.% | 47 (19%) | 36 (17%) | 11 (26%) | 0.2 |
| Pneumonia-no.% | 95 (38%) | 72 (34%) | 23 (52%) | **0.019** |
| Nontuberculous mycobacteria-no.% | 25 (9.9%) | 17 (8.1%) | 8 (19%) | **0.049** |
| C.difficile-no.% | 24 (9.5%) | 18 (8.6%) | 6 (14%) | 0.3 |
| Other infections-no.% | 125 (50%) | 103 (49%) | 22 (51%) | 0.8 |
| COVID-19-no.% | 113 (45%) | 92 (44%) | 21 (49%) | 0.6 |
| Mortality-no.% | 61 (24%) | 48 (23%) | 13 (32%) | 0.3 |
| *^1^* n (%)  *^2^* Pearson’s Chi-squared test; Fisher’s exact test  Bolded p-values indicates statistical significance.  CAV, cardiac allograft vasculopathy; DSA, donor-specific antibody; CMV, Cytomegalovirus; EBV, Epstein–Barr virus; UTI, urinary tract infection; C.difficle, Clostridioides difficile; OHT, orthotopic heart transplant. | | | | |

### Detailed Profile of CH Mutations and Characteristics of Study Cohort Stratified by CH Status

##### **Supplementary Table 3.** Detailed profile of CH mutations identified in the OHT study cohort.

| **Study ID** | **Gene** | **Chromo-**  **some** | **Position** | **Ref Allele** | **Alt Allele** | **Variant Type** | **Variant Type** | **Protein Change** | **Non Reference Read Depth** | **Total Depth** | **VAF** |
| --- | --- | --- | --- | --- | --- | --- | --- | --- | --- | --- | --- |
| SOT_1007 | ASXL1 | chr20 | 31023710 | G | T | missense_variant | SNV | p.Trp1065Cys | 581 | 26528 | 0.021901387 |
| SOT_1008 | ASXL1 | chr20 | 31022234 | G | A | splice_acceptor_variant | INDEL | NM_015338.6:c.1720-1G>A | 1581 | 31622 | 0.049996838 |
| SOT_1020 | SF3B1 | chr2 | 198266779 | G | T | missense_variant | SNV | p.Pro718His | 489 | 16011 | 0.030541503 |
| SOT_1022 | TP53 | chr17 | 7577120 | C | G | missense_variant | SNV | p.Arg273Pro | 5201 | 15698 | 0.331316091 |
| SOT_1026 | DNMT3A | chr2 | 25469104 | G | A | missense_variant | SNV | p.Pro452Ser | 850 | 12289 | 0.069167548 |
| SOT_1029 | U2AF1 | chr21 | 44514777 | T | G | missense_variant | SNV | p.Gln157Pro | 3428 | 15169 | 0.225987211 |
| SOT_1031 | DNMT3A | chr2 | 25458591 | C | T | missense_variant | SNV | p.Cys861Tyr | 815 | 20117 | 0.040512999 |
| SOT_1033 | TET2 | chr4 | 106196618 | C | A | missense_variant | SNV | p.Pro1651Thr | 1164 | 22790 | 0.051075033 |
| SOT_1043 | DNMT3A | chr2 | 25457243 | G | A | missense_variant | SNV | p.Arg882Cys | 6339 | 65424 | 0.096891049 |
| SOT_1052 | ASXL1 | chr20 | 31021206 | G | A | missense_variant | SNV | p.Arg402Gln | 8714 | 19479 | 0.44735356 |
| SOT_1058 | NRAS | chr1 | 115258718 | G | T | missense_variant | SNV | p.Gln22Lys | 1126 | 39502 | 0.028504886 |
| SOT_1079 | ASXL1 | chr20 | 31023679 | G | T | missense_variant | SNV | p.Arg1055Met | 919 | 33887 | 0.027119544 |
| SOT_1086 | IDH2 | chr15 | 90631946 | T | C | missense_variant | SNV | p.Asn136Ser | 16838 | 40884 | 0.411848156 |
| SOT_1092 | RUNX1 | chr21 | 36171741 | G | T | missense_variant | SNV | p.Pro275Gln | 759 | 34419 | 0.022051774 |
| SOT_1094 | DNMT3A | chr2 | 25458637 | G | T | missense_variant | SNV | p.Gln846Lys | 863 | 22891 | 0.037700406 |
| SOT_1094 | KRAS | chr12 | 25398281 | C | T | missense_variant | SNV | p.Gly13Asp | 575 | 22436 | 0.025628454 |
| SOT_1110 | DNMT3A | chr2 | 25463187 | A | G | missense_variant | SNV | p.Ile769Thr | 3646 | 41336 | 0.088203987 |
| SOT_1111 | PHF6 | chrX | 133511739 | T | + | frameshift_variant | INDEL | p.Leu31Phefs*5 | 16968 | 394339 | 0.043028967 |
| SOT_1113 | TET2 | chr4 | 106193796 | T | - | frameshift_variant | INDEL | p.Leu1420Serfs*3 | 10393 | 38739 | 0.268282609 |
| SOT_1113 | TET2 | chr4 | 106193797 | T | - | n/a | INDEL | - | 10393 | 38739 | 0.268282609 |
| SOT_1113 | TET2 | chr4 | 106193798 | A | - | n/a | INDEL | - | 10393 | 38739 | 0.268282609 |
| SOT_1113 | TET2 | chr4 | 106193799 | T | - | n/a | INDEL | - | 10394 | 38739 | 0.268308423 |
| SOT_1113 | TET2 | chr4 | 106193800 | A | - | n/a | INDEL | - | 10394 | 38739 | 0.268308423 |
| SOT_1113 | TET2 | chr4 | 106193801 | C | - | n/a | INDEL | - | 10394 | 38739 | 0.268308423 |
| SOT_1113 | TET2 | chr4 | 106193802 | A | - | n/a | INDEL | - | 10393 | 38739 | 0.268282609 |
| SOT_1113 | TET2 | chr4 | 106193803 | A | - | n/a | INDEL | - | 10380 | 38739 | 0.26794703 |
| SOT_1115 | JAK2 | chr9 | 5073739 | C | A | missense_variant | SNV | p.His606Gln | 23123 | 55992 | 0.41296971 |
| SOT_1123 | PTPN11 | chr12 | 112915487 | G | T | missense_variant | SNV | p.Asp296Tyr | 1237 | 53228 | 0.023239648 |
| SOT_1129 | TET2 | chr4 | 106155397 | G | T | stop_gained | SNV | p.Glu100* | 456 | 16674 | 0.027347967 |
| SOT_1153 | PHF6 | chrX | 133551295 | G | T | missense_variant | SNV | p.Ala311Ser | 875 | 17034 | 0.051367853 |
| SOT_1155 | SMC1A | chrX | 53432021 | C | A | stop_gained | SNV | p.Gly707* | 1341 | 26917 | 0.049819816 |
| SOT_1159 | DNMT3A | chr2 | 25463588 | C | G | missense_variant | SNV | p.Trp698Cys | 6615 | 18475 | 0.358051421 |
| SOT_1322 | FLT3 | chr13 | 28609763 | C | A | missense_variant | SNV | p.Arg489Ile | 1784 | 53644 | 0.033256282 |
| SOT_1325 | TET2 | chr4 | 106193787 | G | T | missense_variant | SNV | p.Val1417Phe | 698 | 18419 | 0.037895651 |
| SOT_1325 | DNMT3A | chr2 | 25466811 | C | G | missense_variant | SNV | p.Arg631Thr | 637 | 6145 | 0.103661513 |
| SOT_1325 | TET2 | chr4 | 106197036 | T | + | frameshift_variant | INDEL | p.Ser1791Phefs*5 | 2068 | 50510 | 0.040942388 |
| SOT_1329 | DNMT3A | chr2 | 25464537 | C | T | missense_variant | SNV | p.Arg659His | 3646 | 47715 | 0.07641203 |
| SOT_1329 | TET2 | chr4 | 106196521 | C | + | frameshift_variant | INDEL | p.Gly1620Trpfs*41 | 1426 | 56569 | 0.025208153 |
| SOT_1335 | STAG2 | chrX | 123184107 | C | A | missense_variant | SNV | p.Ala322Asp | 2158 | 43239 | 0.049908647 |
| SOT_1335 | SMC1A | chrX | 53432008 | C | T | missense_variant | SNV | p.Arg711Gln | 4116 | 15295 | 0.269107551 |
| SOT_1339 | PPM1D | chr17 | 58740542 | A | - | stop_gained | INDEL | p.Thr483* | 10242 | 48802 | 0.209868448 |
| SOT_1339 | PPM1D | chr17 | 58740543 | C | - | n/a | INDEL | - | 10234 | 48803 | 0.209700223 |
| SOT_1339 | PPM1D | chr17 | 58740544 | T | - | n/a | INDEL | - | 10234 | 48804 | 0.209695927 |
| SOT_1339 | PPM1D | chr17 | 58740545 | T | - | n/a | INDEL | - | 10235 | 48804 | 0.209716417 |
| SOT_1349 | RAD21 | chr8 | 117866692 | G | T | missense_variant | SNV | p.Ala318Asp | 1535 | 75648 | 0.020291349 |

##### **Supplementary Table 4.** Additional baseline characteristics of OHT recipients according to CH status with a VAF ≥ 2%.

| **Characteristics** | **All,**  (N=209)^1^ | **No CH,**  (N=180)^1^ | **CH,**  (N=29)^1^ | **p-value^2^** |
| --- | --- | --- | --- | --- |
| CMV Match Status |  |  |  | >0.9 |
| CMV negative-no.% | 39 (19%) | 33 (19%) | 6 (21%) |  |
| CMV positive-no.% | 46 (23%) | 39 (22%) | 7 (24%) |  |
| CMV mismatched-no.% | 119 (58%) | 103 (59%) | 16 (55%) |  |
| EBV Match Status |  |  |  | 0.8 |
| EBV negative-no.% | 2 (1.1%) | 2 (1.2%) | 0 (0%) |  |
| EBV positive-no.% | 155 (82%) | 134 (83%) | 21 (81%) |  |
| EBV mismatched-no.% | 31 (16%) | 26 (16%) | 5 (19%) |  |
| HLA Class I only-no.% | 55 (26%) | 46 (26%) | 9 (31%) | 0.5 |
| HLA Class II only-no.% | 25 (12%) | 25 (14%) | 0 (0%) | **0.029** |
| HLA Class I + II-no.% | 27 (13%) | 23 (13%) | 4 (14%) | 0.8 |
| Bypass Time (minutes) | 148 (119, 174) | 143 (118, 171) | 164 (127, 184) | 0.11 |
| CH, Clonal hematopoiesis; CMV, Cytomegalovirus; EBV, Epstein–Barr virus; HLA, human leukocyte antigen; VAF, variant allele frequency; OHT, orthotopic heart transplant  ^1^ Median (Q1,Q3); n (%)  ^2^ Wilcoxon rank sum test; Pearson’s Chi-squared test; Fisher’s exact test  Bolded p-values indicates statistical significance. | | | | |

##### **Supplementary Table 5.** Immunosuppressive medications of OHT recipients according to CH status with a VAF ≥ 2%.

| **Medications** | **All**  (N=209)^1^ | **No CH**  (N=180)^1^ | **CH**  (N=29)^1^ | **p-value^2^** |
| --- | --- | --- | --- | --- |
| **Immunosuppressive therapy at discharge** | | | | |
| Tacrolimus-no.% | 181 (96%) | 157 (97%) | 24 (89%) | 0.089 |
| Mycophenolate-no.% | 186 (98%) | 160 (99%) | 26 (96%) | 0.4 |
| Cyclosporine-no.% | 7 (3.7%) | 5 (3.1%) | 2 (7.4%) | 0.3 |
| Azathioprine-no.% | 1 (0.5%) | 0 (0%) | 1 (3.7%) | 0.14 |
| Sirolimus-no.% | 1 (0.5%) | 0 (0%) | 1 (3.7%) | 0.14 |
| Prednisone-no.% | 189 (100%) | 162 (100%) | 27 (100%) |  |
| Prednisone dose | 30 (25, 35) | 30 (25, 35) | 30 (25, 40) | >0.9 |
| **Other Medications at Discharge** | | | | |
| Statins-no.% | 176 (93%) | 155 (96%) | 21 (78%) | **0.004** |
| Non-statin lipid lowering-no.% | 3 (1.6%) | 3 (1.9%) | 0 (0%) | >0.9 |
| Calcium-channel blocker-no.% | 26 (14%) | 22 (14%) | 4 (15%) | 0.8 |
| Beta-blocker-no.% | 9 (4.8%) | 8 (4.9%) | 1 (3.7%) | >0.9 |
| ACEI/ARB-no.% | 7 (3.7%) | 7 (4.3%) | 0 (0%) | 0.6 |
| Metformin-no.% | 14 (7.4%) | 14 (8.6%) | 0 (0%) | 0.2 |
| **Immunosuppressive therapy at 1 year** | | | | |
| Tacrolimus-no.% | 167 (93%) | 145 (94%) | 22 (88%) | 0.4 |
| Mycophenolate-no.% | 158 (88%) | 136 (88%) | 22 (88%) | >0.9 |
| Cyclosporine-no.% | 11 (6.1%) | 9 (5.8%) | 2 (8.0%) | 0.7 |
| Azathioprine-no.% | 2 (1.1%) | 2 (1.3%) | 0 (0%) | >0.9 |
| Sirolimus-no.% | 17 (9.5%) | 15 (9.7%) | 2 (8.0%) | >0.9 |
| Prednisone-no.% | 149 (83%) | 127 (82%) | 22 (88%) | 0.8 |
| *^1^* n (%); Median (Q1, Q3)  *^2^* Fisher’s exact test; Wilcoxon rank sum test  Bolded p-values indicates statistical significance. | | | | |

##### **Supplementary Table 6.** Laboratory values of OHT recipients according to CH status with a VAF ≥ 2%.

| **Laboratory Values** | **All**  (N=209)^1^ | **No CH**  (N=180)^1^ | **CH**  (N=29)^1^ | **p-value^2^** |
| --- | --- | --- | --- | --- |
| INR (ratio) | 1.73 (1.10, 2.45) | 1.76 (1.10, 2.50) | 1.54 (1.20, 2.10) | 0.6 |
| Creatinine (umol/L) | 118 (91, 157) | 118 (90, 157) | 127 (111, 151) | 0.4 |
| Sodium (umol/L) | 137 (134, 139) | 137 (134, 139) | 136 (134, 138) | 0.4 |
| Potassium (mmol/L) | 4.10 (3.70, 4.40) | 4.10 (3.70, 4.40) | 4.20 (3.80, 4.50) | 0.4 |
| Random glucose (mmol/L) | 5.40 (4.90, 6.40) | 5.40 (4.90, 6.50) | 5.00 (4.50, 5.60) | **0.046** |
| Calcium (mmol/L) | 2.33 (2.19, 2.42) | 2.33 (2.19, 2.41) | 2.35 (2.17, 2.44) | 0.7 |
| Phosphate (mmol/L) | 1.07 (0.92, 1.23) | 1.07 (0.90, 1.23) | 1.08 (0.99, 1.29) | 0.3 |
| Magnesium (mmol/L) | 0.81 (0.69, 0.91) | 0.81 (0.69, 0.92) | 0.80 (0.71, 0.86) | 0.7 |
| Serum Total Bilirubin (umol/L) | 12 (8, 19) | 11 (8, 19) | 15 (8, 17) | 0.5 |
| Hemoglobin (g/L) | 107 (90, 122) | 107 (90, 122) | 113 (87, 123) | >0.9 |
| Hematocrit | 0.33 (0.28, 0.37) | 0.33 (0.28, 0.37) | 0.34 (0.27, 0.39) | 0.8 |
| White blood cell count (x10e9.L) | 7.7 (5.8, 10.5) | 7.8 (5.7, 10.7) | 7.4 (6.1, 10.0) | 0.8 |
| Neutrophils | 5.7 (3.8, 8.5) | 5.7 (3.8, 8.6) | 5.7 (4.2, 7.5) | >0.9 |
| Lymphocytes | 0.91 (0.54, 1.50) | 0.90 (0.54, 1.50) | 1.03 (0.54, 1.60) | 0.6 |
| Monocytes | 0.60 (0.40, 0.90) | 0.60 (0.40, 0.90) | 0.64 (0.44, 0.86) | 0.6 |
| Platelet (x10e9.L) | 206 (166, 260) | 207 (173, 265) | 191 (148, 232) | 0.10 |
| Serum alkaline phosphatase (IU/L) | 87 (62, 120) | 87 (62, 124) | 79 (57, 108) | 0.3 |
| Aspartate Aminotransferase (SGOT) (IU/L) | 26 (19, 34) | 26 (19, 34) | 24 (14, 35) | 0.4 |
| Alanine transaminase (SGPT) (IU/L) | 25 (16, 47) | 24 (16, 48) | 26 (17, 35) | >0.9 |
| CH, Clonal hematopoiesis; OHT, orthotopic heart transplant  ^1^ Median (Q1, Q3)  ^2^ Wilcoxon rank sum test | | | | |

##### **Supplementary Table 7.** Infections of OHT recipients with CH-related mutations with a VAF ≥ 2% compared to those without CH mutations.

| **Post-transplant outcomes** | **All,**  N=209*^1^* | **No CH**,  N = 180*^1^* | **CH**,  N = 29*^1^* | **p-value***^2^* |
| --- | --- | --- | --- | --- |
| Any infection-no.% | 161 (77%) | 138 (77%) | 23 (79%) | 0.8 |
| Sepsis-no.% | 42 (20%) | 39 (22%) | 3 (10%) | 0.2 |
| CMV Viremia-no.% | 44 (21%) | 39 (22%) | 5 (17%) | 0.6 |
| EBV Viremia-no.% | 20 (9.6%) | 16 (8.9%) | 4 (14%) | 0.5 |
| Candida-no.% | 19 (9.1%) | 16 (8.9%) | 3 (10%) | 0.7 |
| Fungal-no.% | 18 (8.7%) | 17 (9.4%) | 1 (3.4%) | 0.5 |
| UTI-no.% | 36 (17%) | 29 (16%) | 7 (24%) | 0.3 |
| Pneumonia-no.% | 72 (34%) | 64 (36%) | 8 (28%) | 0.4 |
| Nontuberculous mycobacteria-no.% | 17 (8.1%) | 17 (9.4%) | 0 (0%) | 0.14 |
| C.difficile-no.% | 18 (8.6%) | 15 (8.3%) | 3 (10%) | 0.7 |
| Other infections-no.% | 103 (49%) | 87 (48%) | 16 (55%) | 0.5 |
| COVID-19-no.% | 92 (44%) | 77 (43%) | 15 (52%) | 0.4 |
| CH, Clonal hematopoiesis; CMV, Cytomegalovirus; EBV, Epstein–Barr virus; UTI, urinary tract infection; C.difficle, Clostridioides difficile.  ^1^n (%). ^2^Pearson’s Chi-squared test; Fisher’s exact test  Bolded p-values indicates statistical significance. | | | | |

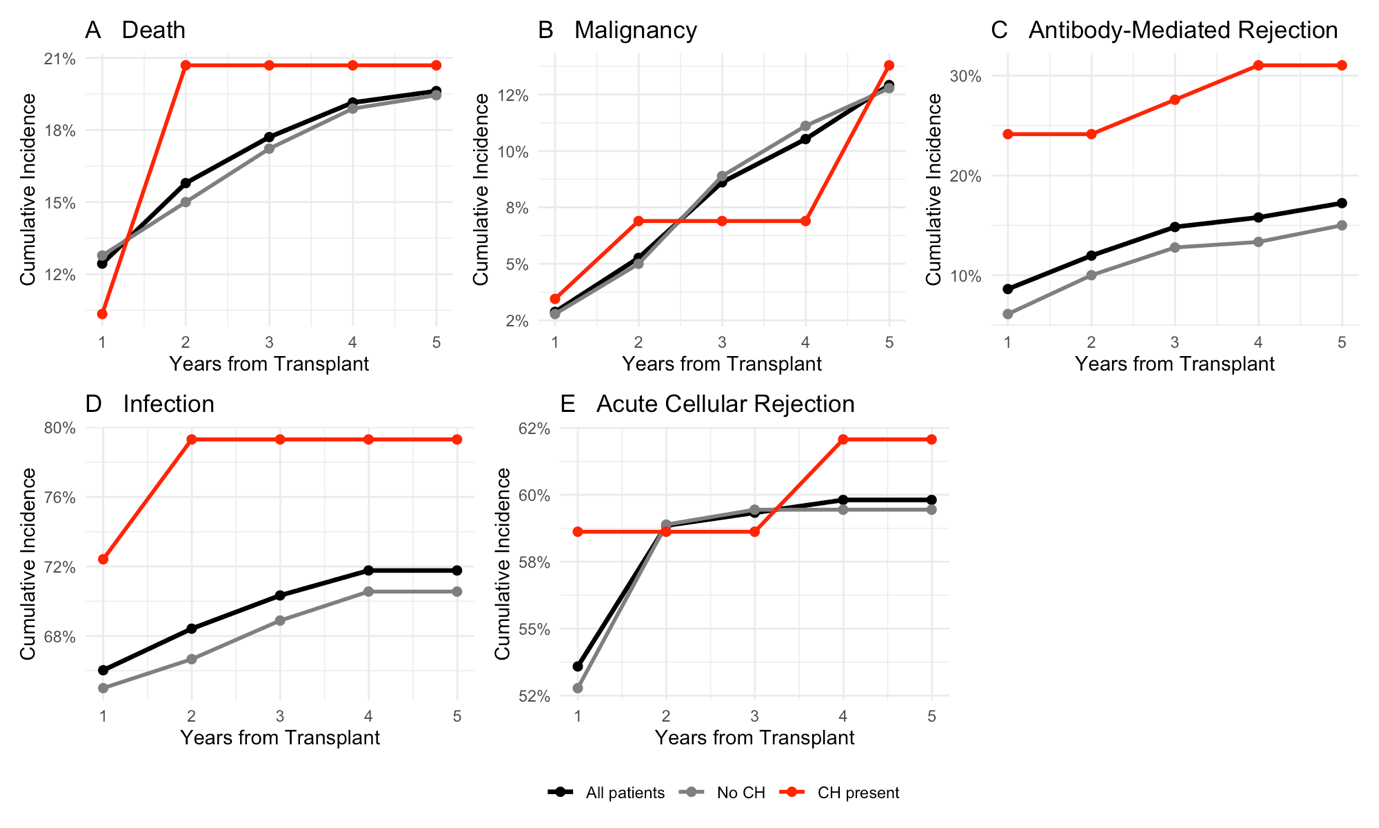

##### **Supplementary Figure 2.** Five-year cumulative incidence of major post-transplant outcomes stratified by CH status with a VAF ≥ 2%.

Panel A represents all-cause mortality. Panel B displays malignancy. Panel C shows antibody-mediated rejection (AMR) Grade 1-3. Panel D represents first incident of infection documented. Finally, Panel E displays acute cellular rejection (ACR) – moderate or severe ACR (≥ 2R). VAF, variant allele frequency; CH, clonal hematopoiesis.

##### **Supplementary Table 8.** Cause of death of OHT recipients with CH-related mutations with a VAF ≥ 2% compared to those without CH mutations.

| **Characteristic** | **No CH,** N = 42*^1^* | **CH,** N = 6*^1^* | **p-value***^2^* |
| --- | --- | --- | --- |
| Graft-Related/Cardiac-no.% | 13 (31%) | 3 (50%) | 0.4 |
| Infection-no.% | 7 (17%) | 2 (33%) | 0.3 |
| Malignancy-no.% | 3 (7.1%) | 1 (17%) | 0.4 |
| Multi-Organ System Failure-no.% | 10 (24%) | 0 (0%) | 0.3 |
| Other-no.% | 3 (7.1%) | 0 (0%) | >0.9 |
| Unknown-no.% | 6 (14%) | 0 (0%) | >0.9 |
| *^1^* n (%) | | | |
| *^2^* Fisher’s exact test | | | |

### Comparison of OHT Recipients by Timing of Biospecimen Collection (±6 Months of Transplant)

##### **Supplementary Table 9.** Post-transplant outcomes of patients who had a biospecimen collected pre-transplant (−6 months to the day before surgery), stratified by CH Status with a VAF ≥ 2%.

This table compares post-transplant outcomes of patients with biospecimens collected before transplant (within 6 months prior to the surgery date), stratified by presence or absence of CH. The "All Patients" column summarizes all patients with a biospecimen 6-month pre-transplant.

| **Characteristic** | **All Patients,**  **N = 112^1^** | **No CH,** n = 96*^1^* | **CH,** n = 16*^1^* | **p-value***^2^* |
| --- | --- | --- | --- | --- |
| Antibody-Mediated Rejection Grade 1-3-no.% | 15 (13%) | 10 (10%) | 5 (31%) | **0.039** |
| CAV Grade |  |  |  | >0.9 |
| CAV 0/1-no.% | 108 (96%) | 92 (96%) | 16 (100%) |  |
| CAV 2/3-no.% | 4 (3.6%) | 4 (4.2%) | 0 (0%) |  |
| Acute cellular rejection (2/3R)-no.% | 57 (51%) | 49 (51%) | 8 (50%) | >0.9 |
| Acute cellular rejection-first-year post-transplant-no.% | 53 (47%) | 46 (48%) | 7 (44%) | 0.8 |
| De novo DSA-no.% | 17 (15%) | 14 (15%) | 3 (19%) | 0.7 |
| Malignancy-no.% | 19 (17%) | 18 (19%) | 1 (6.3%) | 0.3 |
| Infections |  |  |  |  |
| Any infection-no.% | 79 (71%) | 67 (70%) | 12 (75%) | 0.8 |
| Sepsis-no.% | 20 (18%) | 19 (20%) | 1 (6.3%) | 0.3 |
| CMV Viremia-no.% | 13 (12%) | 12 (13%) | 1 (6.3%) | 0.7 |
| EBV Viremia-no.% | 11 (9.8%) | 9 (9.4%) | 2 (13%) | 0.7 |
| Candida-no.% | 10 (8.9%) | 8 (8.3%) | 2 (13%) | 0.6 |
| Fungal-no.% | 10 (8.9%) | 10 (10%) | 0 (0%) | 0.4 |
| UTI-no.% | 17 (15%) | 14 (15%) | 3 (19%) | 0.7 |
| Pneumonia-no.% | 39 (35%) | 34 (35%) | 5 (31%) | 0.7 |
| Nontuberculous mycobacteria-no.% | 10 (8.9%) | 10 (10%) | 0 (0%) | 0.4 |
| C.difficile-no.% | 10 (8.9%) | 7 (7.3%) | 3 (19%) | 0.2 |
| Other infections-no.% | 49 (44%) | 39 (41%) | 10 (63%) | 0.10 |
| COVID-19-no.% | 44 (39%) | 35 (36%) | 9 (56%) | 0.13 |
| Mortality-no.% | 29 (26%) | 27 (28%) | 2 (13%) | 0.2 |
| *^1^* n (%)  *^2^* Pearson’s Chi-squared test; Fisher’s exact test  Bolded p-values indicates statistical significance.  CAV, cardiac allograft vasculopathy; DSA, donor-specific antibody; CMV, Cytomegalovirus; EBV, Epstein–Barr virus; UTI, urinary tract infection; C.difficle, Clostridioides difficile; OHT, orthotopic heart transplant. | | | | |

##### **Supplementary Table 10.** Post-transplant outcomes of OHT recipients with a biospecimen collected from day of transplant to 6 months post-transplant, stratified by CH status with a VAF ≥ 2%.

This table compares post-transplant outcomes among OHT recipients whose biospecimens were collected from the day of transplant (day 0) to 6 months post-transplant. Patients are stratified based on the presence or absence of CH detected in these post-transplant samples. The "All Patients" column summarizes the overall cohort within this biospecimen collection window.

| **Characteristic** | **All Patients,**  **N = 97^1^** | **No CH,** n = 84*^1^* | **CH,** n = 13*^1^* | **p-value***^2^* |
| --- | --- | --- | --- | --- |
| Antibody-Mediated Rejection Grade 1-3-no.% | 22 (23%) | 18 (21%) | 4 (31%) | 0.5 |
| CAV Grade |  |  |  | 0.2 |
| CAV 0/1-no.% | 90 (93%) | 79 (94%) | 11 (85%) |  |
| CAV 2/3-no.% | 7 (7.2%) | 5 (6.0%) | 2 (15%) |  |
| Acute cellular rejection (2/3R)-no.% | 69 (71%) | 59 (70%) | 10 (77%) | 0.8 |
| Acute cellular rejection-first-year post-transplant-no.% | 60 (62%) | 50 (60%) | 10 (77%) | 0.4 |
| De novo DSA-no.% | 21 (22%) | 18 (22%) | 3 (23%) | >0.9 |
| Malignancy-no.% | 20 (21%) | 16 (19%) | 4 (31%) | 0.5 |
| Infections |  |  |  |  |
| Any infection-no.% | 82 (85%) | 71 (85%) | 11 (85%) | >0.9 |
| Sepsis-no.% | 22 (23%) | 20 (24%) | 2 (15%) | 0.7 |
| CMV Viremia-no.% | 31 (32%) | 27 (32%) | 4 (31%) | >0.9 |
| EBV Viremia-no.% | 9 (9.3%) | 7 (8.3%) | 2 (15%) | 0.3 |
| Candida-no.% | 9 (9.3%) | 8 (9.5%) | 1 (7.7%) | >0.9 |
| Fungal-no.% | 8 (8.2%) | 7 (8.3%) | 1 (7.7%) | >0.9 |
| UTI-no.% | 19 (20%) | 15 (18%) | 4 (31%) | 0.3 |
| Pneumonia-no.% | 33 (34%) | 30 (36%) | 3 (23%) | 0.5 |
| Nontuberculous mycobacteria-no.% | 7 (7.2%) | 7 (8.3%) | 0 (0%) | 0.6 |
| C.difficile-no.% | 8 (8.2%) | 8 (9.5%) | 0 (0%) | 0.6 |
| Other Infections-no.% | 54 (56%) | 48 (57%) | 6 (46%) | 0.5 |
| COVID-19-no.% | 48 (49%) | 42 (50%) | 6 (46%) | 0.8 |
| Mortality-no.% | 19 (20%) | 15 (18%) | 4 (31%) | 0.3 |
| *^1^* n (%)  *^2^* Pearson’s Chi-squared test; Fisher’s exact test  CAV, cardiac allograft vasculopathy; DSA, donor-specific antibody; CMV, Cytomegalovirus; EBV, Epstein–Barr virus; UTI, urinary tract infection; C.difficle, Clostridioides difficile; OHT,  orthotopic heart transplant. | | | | |

### Gene Specific Analysis (Epigenetic Regulator Genes)

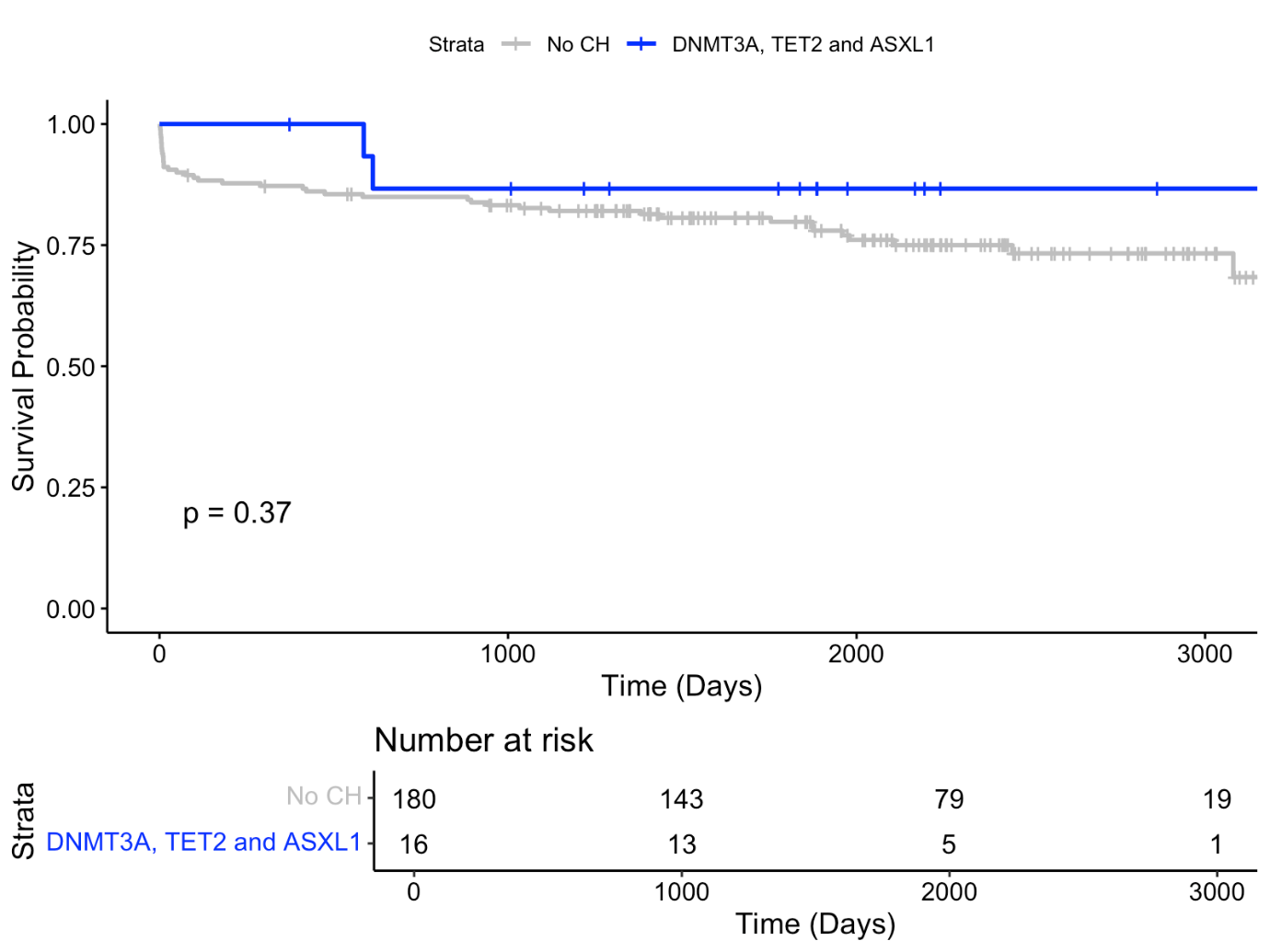

##### **Supplementary Figure 3.** Kaplan-Meier survival curve of OHT recipients stratified by DNMT3A, TET2 and ASXL1 mutation status with a VAF ≥ 2%.

##### **Supplementary Table 11.** Baseline characteristics of OHT recipients with CH-related mutations in DNMT3A, TET2, and ASXL1 with a VAF ≥ 2% compared to those without CH mutations.

| **Characteristic** | **No CH,** n = 180*^1^* | **DNMT3A, TET2 and ASXL1 mutations,** n = 16*^1^* | **p-value***^2^* |
| --- | --- | --- | --- |
| Age (years) | 53 (41, 61) | 51 (46, 58) | 0.8 |
| Sex |  |  | 0.4 |
| Male-no. (%) | 133 (74%) | 10 (63%) |  |
| Female-no. (%) | 47 (26%) | 6 (38%) |  |
| BMI (kg/m^2^) | 25.4 (22.3, 28.9) | 23.7 (22.7, 29.4) | 0.7 |
| History of dyslipidemia-no. (%) | 47 (26%) | 4 (25%) | >0.9 |
| History of diabetes mellitus-no.% | 39 (22%) | 1 (6.3%) | 0.2 |
| History of hypertension-no.% | 47 (26%) | 2 (13%) | 0.4 |
| History of myocardial infarction-no.% | 31 (17%) | 0 (0%) | 0.081 |
| Previous PCI -no.% | 22 (12%) | 0 (0%) | 0.2 |
| Previous CABG-no.% | 11 (6.1%) | 0 (0%) | 0.6 |
| History of CVA/TIA-no.% | 31 (17%) | 5 (31%) | 0.2 |
| History of atrial fibrillation/flutter-no.% | 52 (29%) | 5 (31%) | 0.8 |
| History of CKD-no.% | 53 (29%) | 4 (25%) | >0.9 |
| LVAD-no.% | 59 (33%) | 4 (25%) | 0.5 |
| History of alcohol abuse-no.% | 11 (6.1%) | 1 (6.3%) | >0.9 |
| History of smoking-no.% | 46 (26%) | 3 (19%) | 0.8 |
| Intracardiac device |  |  | 0.2 |
| ICD-no.% | 69 (50%) | 8 (62%) |  |
| CRT-P-no.% | 3 (2.2%) | 1 (7.7%) |  |
| CRT-D-no.% | 66 (48%) | 4 (31%) |  |
| Etiology of Heart Failure |  |  | 0.3 |
| Ischemic-no.% | 39 (22%) | 2 (13%) |  |
| Non-ischemic-no.% | 120 (67%) | 10 (63%) |  |
| Congenital-no.% | 21 (12%) | 4 (25%) |  |
| ABO Group |  |  | 0.7 |
| A-no.% | 68 (38%) | 8 (50%) |  |
| B-no.% | 25 (14%) | 2 (13%) |  |
| AB-no.% | 10 (5.6%) | 1 (6.3%) |  |
| O-no.% | 77 (43%) | 5 (31%) |  |
| Bypass Time (minutes) | 143 (118, 171) | 164 (124, 180) | 0.2 |
| Total ischemic time (minutes) | 204 (163, 248) | 200 (154, 207) | 0.4 |
| CMV Match Status |  |  | 0.6 |
| CMV negative-no.% | 33 (19%) | 3 (19%) |  |
| CMV positive-no.% | 39 (22%) | 5 (31%) |  |
| CMV mismatched-no.% | 103 (59%) | 8 (50%) |  |
| EBV Match Status |  |  | 0.8 |
| EBV negative-no.% | 2 (1.2%) | 0 (0%) |  |
| EBV positive-no.% | 134 (83%) | 11 (79%) |  |
| EBV mismatched-no.% | 26 (16%) | 3 (21%) |  |
| HLA Class I only-no.% | 46 (26%) | 6 (38%) | 0.4 |
| HLA Class II only-no.% | 25 (14%) | 0 (0%) | 0.2 |
| HLA Class I + II-no.% | 23 (13%) | 3 (19%) | 0.5 |
| *^1^* Median (Q1, Q3); n (%)  *^2^* Wilcoxon rank sum test; Pearson’s Chi-squared test; Fisher’s exact test  BMI, Body mass index; PCI, Percutaneous coronary intervention; CABG, Coronary artery bypass surgery; CVA, cerebral vascular accident; TIA, transient ischemic attack; CKD, chronic kidney disease; LVAD, left ventricular assist device; ICD, implantable cardioverter-defibrillator; CRT-D, cardiac resynchronization therapy (CRT) defibrillator; CRT-P, cardiac resynchronization therapy pacemaker; CMV, Cytomegalovirus; EBV, Epstein–Barr virus; HLA, human leukocyte antigen; VAF, variant allele frequency; OHT, orthotopic heart transplant | | | |

##### **Supplementary Table 12.** Early post-transplant outcomes of OHT recipients according to mutations in DNMT3A, TET2 and ASXL1 with a VAF ≥ 2%.

| **Early post-transplant outcomes** | **No CH,** n = 180*^1^* | **DNMT3A, TET2 and ASXL1 mutations,** n = 16*^1^* | **p-value***^2^* |
| --- | --- | --- | --- |
| Primary graft dysfunction-no.% | 42 (24%) | 3 (20%) | >0.9 |
| Need temporary circulatory mechanical support-no.% | 18 (10%) | 3 (20%) | 0.2 |
| Need for temporary dialysis-no.% | 18 (10%) | 1 (6.7%) | >0.9 |
| *^1^* n (%)  *^2^*  Fisher’s exact test | | | |

##### **Supplementary Table 13.** Post-transplant outcomes of OHT recipients with CH-related mutations in DNMT3A, TET2 and ASXL1 with a VAF ≥ 2% compared to those without CH mutations.

| **Characteristic** | **No CH,** n = 180*^1^* | **DNMT3A, TET2 and ASXL1 mutations,** n = 16*^1^* | **p-value***^2^* |
| --- | --- | --- | --- |
| Antibody-Mediated Rejection Grade 1-3-no.% | 28 (16%) | 2 (13%) | >0.9 |
| CAV Grade |  |  | >0.9 |
| CAV 0/1-no.% | 171 (95%) | 16 (100%) |  |
| CAV 2/3-no.% | 9 (5.0%) | 0 (0%) |  |
| Acute cellular rejection (2/3R)-no.% | 108 (60%) | 10 (63%) | 0.8 |
| Acute cellular rejection-first-year post-transplant-no.% | 96 (53%) | 10 (63%) | 0.5 |
| De novo DSA-no.% | 32 (18%) | 4 (25%) | 0.5 |
| Malignancy-no.% | 34 (19%) | 4 (25%) | 0.5 |
| Infections |  |  |  |
| Any infection-no.% | 138 (77%) | 14 (88%) | 0.5 |
| Sepsis-no.% | 39 (22%) | 2 (13%) | 0.5 |
| CMV Viremia-no.% | 39 (22%) | 5 (31%) | 0.4 |
| EBV Viremia-no.% | 16 (8.9%) | 3 (19%) | 0.2 |
| Candida-no.% | 16 (8.9%) | 3 (19%) | 0.2 |
| Fungal-no.% | 17 (9.4%) | 0 (0%) | 0.4 |
| UTI-no.% | 29 (16%) | 1 (6.3%) | 0.5 |
| Pneumonia-no.% | 64 (36%) | 5 (31%) | 0.7 |
| Nontuberculous mycobacteria-no.% | 17 (9.4%) | 0 (0%) | 0.4 |
| C.difficile-no.% | 15 (8.3%) | 2 (13%) | 0.6 |
| Other infections-no.% | 87 (48%) | 9 (56%) | 0.5 |
| COVID-19-no.% | 77 (43%) | 9 (56%) | 0.3 |
| Mortality-no.% | 42 (23%) | 2 (13%) | 0.5 |
| *^1^* n (%)  *^2^* Fisher’s exact test; Pearson’s Chi-squared test  CH, Clonal hematopoiesis; CAV, cardiac allograft vasculopathy; DSA, donor-specific antibody; CMV, Cytomegalovirus; EBV, Epstein–Barr virus; UTI, urinary tract infection; C.difficle, Clostridioides difficile. | | | |

##### **Supplementary Table 14.** Post-transplant outcomes of OHT recipients with CH-related mutations in DNMT3A with a VAF ≥ 2% compared to those without CH mutations.

| **Characteristic** | **No CH,** N = 180*^1^* | **DNMT3A Mutations,** N = 8*^1^* | **p-value***^2^* |
| --- | --- | --- | --- |
| Antibody-Mediated Rejection Grade 1-3-no.% | 28 (16%) | 1 (13%) | >0.9 |
| CAV Grade |  |  | >0.9 |
| CAV 0/1-no.% | 171 (95%) | 8 (100%) |  |
| CAV 2/3-no.% | 9 (5.0%) | 0 (0%) |  |
| Acute cellular rejection (2/3R)-no.% | 108 (60%) | 7 (88%) | 0.2 |
| Acute cellular rejection-first-year post-transplant-no.% | 96 (53%) | 7 (88%) | 0.074 |
| De novo DSA-no.% | 32 (18%) | 3 (38%) | 0.2 |
| Malignancy-no.% | 34 (19%) | 2 (25%) | 0.7 |
| Infections |  |  |  |
| Any infection-no.% | 138 (77%) | 7 (88%) | 0.7 |
| Sepsis-no.% | 39 (22%) | 1 (13%) | >0.9 |
| CMV Viremia-no.% | 39 (22%) | 2 (25%) | 0.7 |
| EBV Viremia-no.% | 16 (8.9%) | 2 (25%) | 0.2 |
| Candida-no.% | 16 (8.9%) | 2 (25%) | 0.2 |
| Fungal-no.% | 17 (9.4%) | 0 (0%) | >0.9 |
| UTI-no.% | 29 (16%) | 1 (13%) | >0.9 |
| Pneumonia-no.% | 64 (36%) | 2 (25%) | 0.7 |
| Nontuberculous mycobacteria-no.% | 17 (9.4%) | 0 (0%) | >0.9 |
| C.difficile-no.% | 15 (8.3%) | 1 (13%) | 0.5 |
| Other infections-no.% | 87 (48%) | 5 (63%) | 0.5 |
| COVID-19-no.% | 77 (43%) | 5 (63%) | 0.3 |
| Mortality-no.% | 42 (23%) | 1 (13%) | 0.7 |
| *^1^* n (%)  *^2^* Fisher’s exact test  Bolded p-values indicates statistical significance.  CH, Clonal hematopoiesis; CAV, cardiac allograft vasculopathy; DSA, donor-specific antibody; CMV, Cytomegalovirus; EBV, Epstein–Barr virus; UTI, urinary tract infection; C.difficle, Clostridioides difficile. | | | |

##### **Supplementary Table 15.** Post-transplant outcomes of OHT recipients with CH-related mutations in TET2 with a VAF ≥ 2% compared to those without CH mutations.

| **Characteristic** | **No CH,** N = 180*^1^* | **TET2 Mutations,** N = 4*^1^* | **p-value***^2^* |
| --- | --- | --- | --- |
| Antibody-Mediated Rejection Grade 1-3-no.% | 28 (16%) | 0 (0%) | >0.9 |
| CAV Grade |  |  | >0.9 |
| CAV 0/1-no.% | 171 (95%) | 4 (100%) |  |
| CAV 2/3-no.% | 9 (5.0%) | 0 (0%) |  |
| Acute cellular rejection (2/3R)-no.% | 108 (60%) | 3 (75%) | >0.9 |
| Acute cellular rejection-first-year post-transplant-no.% | 96 (53%) | 3 (75%) | 0.6 |
| De novo DSA-no.% | 32 (18%) | 0 (0%) | >0.9 |
| Malignancy-no.% | 34 (19%) | 1 (25%) | 0.6 |
| Infections |  |  |  |
| Any infection-no.% | 138 (77%) | 4 (100%) | 0.6 |
| Sepsis-no.% | 39 (22%) | 1 (25%) | >0.9 |
| CMV Viremia-no.% | 39 (22%) | 2 (50%) | 0.2 |
| EBV Viremia-no.% | 16 (8.9%) | 1 (25%) | 0.3 |
| Candida-no.% | 16 (8.9%) | 0 (0%) | >0.9 |
| Fungal-no.% | 17 (9.4%) | 0 (0%) | >0.9 |
| UTI-no.% | 29 (16%) | 0 (0%) | >0.9 |
| Pneumonia-no.% | 64 (36%) | 2 (50%) | 0.6 |
| Nontuberculous mycobacteria-no.% | 17 (9.4%) | 0 (0%) | >0.9 |
| C.difficile-no.% | 15 (8.3%) | 0 (0%) | >0.9 |
| Other infections-no.% | 87 (48%) | 3 (75%) | 0.4 |
| COVID-19-no.% | 77 (43%) | 3 (75%) | 0.3 |
| Mortality-no.% | 42 (23%) | 1 (25%) | >0.9 |
| *^1^* n (%)  *^2^* Fisher’s exact test  Bolded p-values indicates statistical significance.  CH, Clonal hematopoiesis; CAV, cardiac allograft vasculopathy; DSA, donor-specific antibody; CMV, Cytomegalovirus; EBV, Epstein–Barr virus; UTI, urinary tract infection; C.difficle, Clostridioides difficile. | | | |

##### **Supplementary Table 16.** Post-transplant outcomes of OHT recipients with CH-related mutations in ASXL1 with a VAF ≥ 2% compared to those without CH mutations.

| **Characteristic** | **No CH,** N = 180*^1^* | **ASXL1 Mutations,** N = 4*^1^* | **p-value***^2^* |
| --- | --- | --- | --- |
| Antibody-Mediated Rejection Grade 1-3-no.% | 28 (16%) | 1 (25%) | 0.5 |
| CAV Grade |  |  | >0.9 |
| CAV 0/1-no.% | 171 (95%) | 4 (100%) |  |
| CAV 2/3-no.% | 9 (5.0%) | 0 (0%) |  |
| Acute cellular rejection (2/3R)-no.% | 108 (60%) | 0 (0%) | **0.028** |
| Acute cellular rejection-first-year post-transplant-no.% | 96 (53%) | 0 (0%) | **0.050** |
| De novo DSA-no.% | 32 (18%) | 1 (25%) | 0.5 |
| Malignancy-no.% | 34 (19%) | 1 (25%) | 0.6 |
| Infections |  |  |  |
| Any infection-no.% | 138 (77%) | 3 (75%) | >0.9 |
| Sepsis-no.% | 39 (22%) | 0 (0%) | 0.6 |
| CMV Viremia-no.% | 39 (22%) | 1 (25%) | >0.9 |
| EBV Viremia-no.% | 16 (8.9%) | 0 (0%) | >0.9 |
| Candida-no.% | 16 (8.9%) | 1 (25%) | 0.3 |
| Fungal-no.% | 17 (9.4%) | 0 (0%) | >0.9 |
| UTI-no.% | 29 (16%) | 0 (0%) | >0.9 |
| Pneumonia-no.% | 64 (36%) | 1 (25%) | >0.9 |
| Nontuberculous mycobacteria-no.% | 17 (9.4%) | 0 (0%) | >0.9 |
| C.difficile-no.% | 15 (8.3%) | 1 (25%) | 0.3 |
| Other infections-no.% | 87 (48%) | 1 (25%) | 0.6 |
| COVID-19-no.% | 77 (43%) | 1 (25%) | 0.6 |
| Mortality-no.% | 42 (23%) | 0 (0%) | 0.6 |
| *^1^* n (%)  *^2^* Fisher’s exact test  Bolded p-values indicates statistical significance.  CH, Clonal hematopoiesis; CAV, cardiac allograft vasculopathy; DSA, donor-specific antibody; CMV, Cytomegalovirus; EBV, Epstein–Barr virus; UTI, urinary tract infection; C.difficle, Clostridioides difficile. | | | |

##### **Supplementary Table 17.** Multivariate Cox proportional hazards analysis of CAV Grade 1-3 by DNMT3A, TET2 and ASXL1 mutation status with a VAF ≥ 2%.

| **Characteristic** | **HR** | **95% CI** | **p-value** |
| --- | --- | --- | --- |
| DNMT3A, TET2 and ASXL1 Mutations | 1.21 | 0.63, 2.32 | 0.6 |
| Age | 1.00 | 0.99, 1.02 | 0.7 |
| Sex | 0.68 | 0.43, 1.08 | 0.10 |
| Ischemic etiology | 1.11 | 0.70, 1.76 | 0.7 |
| Abbreviations: CI = Confidence Interval, HR = Hazard Ratio | | | |

##### **Supplementary Table 18.** Multivariate Cox proportional hazards analysis of CAV 2/3 by DNMT3A, TET2 and ASXL1 mutation status with a VAF ≥ 2%.

| **Characteristic** | **HR** | **95% CI** | **p-value** |
| --- | --- | --- | --- |
| DNMT3A, TET2 and ASXL1 Mutations | 0.00 | 0.00, Inf | >0.9 |
| Age | 0.98 | 0.93, 1.03 | 0.5 |
| Sex | 0.00 | 0.00, Inf | >0.9 |
| Ischemic etiology | 0.93 | 0.18, 4.77 | >0.9 |
| Abbreviations: CI = Confidence Interval, HR = Hazard Ratio | | | |

##### **Supplementary Table 19.** Multivariate Cox proportional hazards analysis of antibody-mediated rejection by DNMT3A, TET2 and ASXL1 mutation status with a VAF ≥ 2%.

| **Characteristic** | **HR** | **95% CI** | **p-value** |
| --- | --- | --- | --- |
| DNMT3A, TET2 and ASXL1 Mutations | 0.81 | 0.19, 3.41 | 0.8 |
| Age | 0.99 | 0.96, 1.01 | 0.3 |
| Sex | 1.56 | 0.71, 3.42 | 0.3 |
| Ischemic etiology | 1.74 | 0.74, 4.10 | 0.2 |
| Abbreviations: CI = Confidence Interval, HR = Hazard Ratio | | | |
