## Supplementary material for "Timing matters: Detection of clonal hematopoiesis and its association with adverse outcomes in heart transplant recipients": Table 1

**Table 1. Baseline characteristics of OHT recipients according to CH status with a VAF** $\boldsymbol{\geq}$ **2%.**

| Characteristics | All,  (N=209)^1^ | No CH,  (N=180)^1^ | CH,  (N=29)^1^ | p-value^2^ |
| --- | --- | --- | --- | --- |
| Age (years) | 53 (41, 60) | 53 (41, 61) | 51 (41, 58) | 0.5 |
| Sex | | | | 0.6 |
| Male-no. (%) | 153 (73%) | 133 (74%) | 20 (69%) |  |
| Female-no. (%) | 56 (27%) | 47 (26%) | 9 (31%) |  |
| BMI (kg/m^2^) | 25.2 (22.4, 28.9) | 25.4 (22.3, 28.9) | 24.1 (22.7, 30.2) | >0.9 |
| Comorbidities |  |  |  |  |
| History of dyslipidemia-no. (%) | 54 (26%) | 47 (26%) | 7 (24%) | 0.8 |
| History of diabetes mellitus-no.% | 42 (20%) | 39 (22%) | 3 (10%) | 0.2 |
| History of hypertension-no.% | 51 (24%) | 47 (26%) | 4 (14%) | 0.2 |
| History of myocardial infarction-no.% | 33 (16%) | 31 (17%) | 2 (6.9%) | 0.3 |
| Previous PCI-no.% | 24 (11%) | 22 (12%) | 2 (6.9%) | 0.5 |
| Previous CABG-no.% | 12 (5.7%) | 11 (6.1%) | 1 (3.4%) | >0.9 |
| History of CVA/TIA-no.% | 37 (18%) | 31 (17%) | 6 (21%) | 0.6 |
| History of atrial fibrillation/flutter-no.% | 59 (28%) | 52 (29%) | 7 (24%) | 0.6 |
| History of CKD-no.% | 59 (28%) | 53 (29%) | 6 (21%) | 0.3 |
| LVAD-no.% | 67 (32%) | 59 (33%) | 8 (28%) | 0.6 |
| History of alcohol abuse-no.% | 12 (5.7%) | 11 (6.1%) | 1 (3.4%) | >0.9 |
| History of smoking-no.% | 53 (25%) | 46 (26%) | 7 (24%) | 0.9 |
| Intracardiac device | | | | 0.6 |
| ICD-no.% | 81 (50%) | 69 (50%) | 12 (52%) |  |
| CRT-P-no.% | 4 (2.5%) | 3 (2.2%) | 1 (4.3%) |  |
| CRT-D-no.% | 76 (47%) | 66 (48%) | 10 (43%) |  |
| Etiology of Heart Failure | | | | 0.3 |
| Ischemic-no.% | 43 (21%) | 39 (22%) | 4 (14%) |  |
| Non-ischemic-no.% | 139 (67%) | 120 (67%) | 19 (66%) |  |
| Congenital-no.% | 27 (13%) | 21 (12%) | 6 (21%) |  |
| ABO Group | | | | >0.9 |
| A-no.% | 79 (38%) | 68 (38%) | 11 (38%) |  |
| B-no.% | 29 (14%) | 25 (14%) | 4 (14%) |  |
| AB-no.% | 12 (5.7%) | 10 (5.6%) | 2 (6.9%) |  |
| O-no.% | 89 (43%) | 77 (43%) | 12 (41%) |  |
| Total ischemic time (minutes) | 205 (165, 247) | 204 (163, 248) | 205 (174, 227) | 0.8 |
| CH, Clonal hematopoiesis; BMI, Body mass index; PCI, Percutaneous coronary intervention; CABG, Coronary artery bypass surgery; CVA, cerebral vascular accident; TIA, transient ischemic attack; CKD, chronic kidney disease; LVAD, left ventricular assist device; ICD, implantable cardioverter-defibrillator; CRT-D, cardiac resynchronization therapy (CRT) defibrillator; CRT-P, cardiac resynchronization therapy pacemaker; VAF, variant allele frequency; OHT, orthotopic heart transplant  ^1^ Median (Q1,Q3); n (%)  ^2^ Wilcoxon rank sum test; Pearson’s Chi-squared test; Fisher’s exact test | | | | |
