## Supplementary material for "Timing matters: Detection of clonal hematopoiesis and its association with adverse outcomes in heart transplant recipients": Table 2

**Table 2. Post-transplant outcomes of OHT recipients with CH-related mutations with a VAF** $\boldsymbol{\geq}$ **2% compared to those without CH mutations.**

| **Post-transplant outcomes** | **All,**  N=209*^1^* | **No CH**,  N = 180*^1^* | **CH**,  N = 29*^1^* | **p-value***^2^* |
| --- | --- | --- | --- | --- |
| Mortality-no.% | 48 (23%) | 42 (23%) | 6 (21%) | 0.8 |
| Antibody-Mediated Rejection Grade 1-3-no.% | 37 (18%) | 28 (16%) | 9 (31%) | **0.043** |
| Acute cellular rejection (2/3R)-no.% | 126 (60%) | 108 (60%) | 18 (62%) | 0.8 |
| Acute cellular rejection-first-year post-transplant-no.% | 113 (54%) | 96 (53%) | 17 (59%) | 0.6 |
| De novo DSA-no.% | 38 (18%) | 32 (18%) | 6 (21%) | 0.7 |
| Malignancy-no.% | 39 (19%) | 34 (19%) | 5 (17%) | 0.8 |
| CAV grade |  |  |  | 0.7 |
| CAV 0/1-no.% | 198 (95%) | 171 (95%) | 27 (93%) |  |
| CAV 2/3-no.% | 11 (5.3%) | 9 (5.0%) | 2 (6.9%) |  |
| Early post-transplant outcomes | | | | |
| Primary graft dysfunction-no.% | 46 (22%) | 42 (24%) | 4 (14%) | 0.3 |
| Need for temporary mechanical support-no.% | 21 (10%) | 18 (10%) | 3 (11%) | >0.9 |
| Need for temporary dialysis-no.% | 20 (9.7%) | 18 (10%) | 2 (7.1%) | >0.9 |
| CH, Clonal hematopoiesis; CAV, cardiac allograft vasculopathy; DSA, donor-specific antibody.  ^1^ n (%), ^2^ Pearson’s Chi-squared test; Fisher’s exact test  Bolded p-values indicates statistical significance. | | | | |
