## Supplementary material for "Timing matters: Detection of clonal hematopoiesis and its association with adverse outcomes in heart transplant recipients": Table 3

**Table 3. Univariate and multivariate cox proportional hazards analysis of post-transplant outcomes by CH mutation status with a VAF** $\boldsymbol{\geq}$ **2%.**

| Variables | HR*^1^* | 95% CI*^1^* | p-value |
| --- | --- | --- | --- |
| All-cause mortality | | | |
| Univariable^2^ | 0.90 | 0.38, 2.11 | 0.8 |
| Multivariable^3^ | 1.03 | 0.42, 2.52 | >0.9 |
| AMR |  |  |  |
| Univariable^2^ | 2.32 | 1.09, 4.92 | 0.029 |
| Multivariable^3^ | 2.42 | 1.07, 5.47 | 0.033 |
| CAV Grade 1-3 |  |  |  |
| Univariable^2^ | 1.26 | 0.77, 2.06 | 0.4 |
| Multivariable^3^ | 1.46 | 0.85, 2.49 | 0.2 |
| CAV Grade 2/3 |  |  |  |
| Univariable^2^ | 1.43 | 0.31, 6.64 | 0.6 |
| Multivariable^3^ | 1.05 | 0.19, 5.83 | >0.9 |
| ACR |  |  |  |
| Univariable^2^ | 0.97 | 0.59, 1.60 | >0.9 |
| Multivariable^3^ | 0.81 | 0.48, 1.37 | 0.4 |
| Malignancy |  |  |  |
| Univariable^2^ | 0.87 | 0.34, 2.24 | 0.8 |
| Multivariable^3^ | 1.39 | 0.51, 3.75 | 0.5 |
| ^1^ HR = Hazard Ratio, CI = Confidence Interval.  ^2^ Unadjusted.  ^3^ Adjusted for age, sex, BMI, ischemic etiology, diabetes mellitus, hypertension, dyslipidemia, prior myocardial infarction, and pre-transplant intracardiac device implantation.  Bolded p-values indicates statistical significance.  Univariate and multivariate analysis of all-cause mortality, antibody-mediated rejection (AMR), cardiac allograft vasculopathy (CAV), acute cellular rejection (ACR) and malignancy post-transplant. | | | |
