## Supplementary material for "Timing matters: Detection of clonal hematopoiesis and its association with adverse outcomes in heart transplant recipients": Table 4

**Table 4. Summary of studies investigating CH in heart transplant recipients.**

| **Study (Author, Year)** | **Participants** | **CH Prevalence** | **Age at Transplant** | **Sample Timing Relative to Transplant** | **Median follow-up post-OHT** | **CH-Associated Outcomes** | **Top Genes Mutated** |
| --- | --- | --- | --- | --- | --- | --- | --- |
| Our study | 209 | 13.9% | 53 (IQR 41- 60) | Median: 1 day before transplant  Sample collection window: ± 6 months of transplantation | All patients: 5.1 years (IQR 3.3 years)  No CH: 5.1 years (IQR: 3.2 years)  CH: 5.2 years (IQR: 3.6 years) | ↑ AMR (HR 2.42) | DNMT3A, ASXL1, TET2 |
| Scolari et al., 2022 | 127 | 20.5% | 49 ± 14 years | 68% pre-transplant (median ~200 days), 32% post-transplant (median ~114 days) | 3.2 ± 2.6 years | ↑ CAV (18% vs. 0%, p < .001)  ↑ Mortality (42% vs. 15%, adj. HR 2.9) | DNMT3A, ASXL1, TET2 |
| Amancherla et al., 2023 | 479 of 787 included (301 VUMC, 178 CUIMC, 308 VUMC patients excluded despite sequencing) | 14.9% pooled: (13.5% VUMC, 19.7% CUIMC) | Not reported | Median 2.3 years before transplant (VUMC).  Just before transplant (CUIMC) | 18 years (VUMC) or 5.1 years (CUIMC) | No significant association with CAV or mortality  There was a significantly increased risk of CAV with large CHIP clones in the CUIMC cohort, but this was representative of only 6 patients. | DNMT3A, PPM1D, TET2 |
| Simitsis et al., 2025 | 95 | 31.6% | 52 (IQR:18) years  Patients with CHIP mutations were older at the time of enrollment (median age, 71 vs 56 years; P < 0.001) and at the time of transplant (median age, 59 vs 46 years; P < 0.001) | Median 8.5 years after transplant | 9.7 years (IQR: 11.3)  13.3 months (IQR: 4.9) from enrollment | No significant association with CAV, graft failure, malignancy, or mortality | DNMT3A, PPM1D, SF3B1 |
